# Safety and feasibility of blood-derived Multiple Antigen-Specific Endogenous T cells (MASE-T) for metastatic melanoma

**DOI:** 10.1101/2025.07.02.25330108

**Authors:** Tine J. Monberg, Siri Amanda Tvingholm, Marcus Svensson-Frej, Cecilie Vestergaard, Maria Ormhøj, Julie W. Kjeldsen, Troels H. Borch, Rikke B. Holmstroem, Nithiyashri Jayashankar, Joachim S. Granhøj, Anne R. Cordt, Stine K. Larsen, Özcan Met, Sine R. Hadrup, Inge Marie Svane

## Abstract

**Background:** Tumor infiltrating lymphocyte (TIL) therapy is effective in metastatic melanoma, but the need for resectable tumor tissue limits its accessibility. Antigen-presenting scaffolds (Ag-scaffolds) are developed for the specific expansion of tumor-associated antigen (TAA)-specific T cells directly from peripheral blood. Ag-scaffolds are built on a dextran backbone with co-attached interleukin 2 (IL-2), interleukin 21 (IL-21), and MHC I molecules loaded with the top 30 most frequently expressed TAAs in melanoma patients. The resulting multiple antigen-specific endogenously derived T cell (MASE-T) infusion product is characterized by increased CD8+ TAA-specific T cells. We hypothesize that treatment with MASE-T therapy is safe and feasible in patients with immune checkpoint inhibitor (ICI)-resistant metastatic melanoma.

**Methods:** In this phase I, first-in-human, clinical trial, six patients with ICI-resistant melanoma were treated with MASE-T cells preceded by three days of lymphodepleting chemotherapy with cyclophosphamide and fludarabine phosphate.

**Results:** MASE-T cells were successfully expanded in 88% (7/8) of the included patients and most MASE-T products were enriched for T cell populations targeting multiple TAAs.

Administration of MASE-T therapy was safe with no MASE-T-related toxicities. Clinical efficacy was limited, with three out of six (50%) of patients having stable disease six weeks post treatment.

**Conclusions:** This trial demonstrates that Ag-scaffold-driven expansion of TAA-specific T cells from the peripheral blood of patients with melanoma is feasible and the resulting MASE-T infusion product can be safely administered. However, further development is required to unleash the full potential of this technology.

**Trial registration:** Clinicaltrials.gov NCT04904185, registered 22.05.2021.

**What is already known on this topic:** Antigen-presenting scaffolds (Ag-scaffolds) represents a novel artificial antigen presenting cell (aAPC) technology designed to specifically expand tumor-specific T cells from peripheral blood mononuclear cells (PBMCs). Although Ag-scaffold technology has shown promising results in preclinical studies, its application in a large-scale clinical setting remains unexplored.

**What this study adds:** This study demonstrates the clinical applicability of the Ag-scaffold technology in patients with metastatic melanoma. The resulting multiple antigen-specific endogenously derived T cell (MASE-T) infusion product can be safely administered with no MASE-T related adverse events observed.

**How this study might affect research, practice or policy:** This study demonstrates the potential of the Ag-scaffold technology to expand low-frequent CD8+ tumor-specific T cell populations directly from peripheral blood. This proof of concept suggest that the Ag-scaffold technology could serve as a future alternative to treatment with tumor infiltrating lymphocytes (TILs) for patients not eligible for TIL therapy.

## Background

Adoptive cell therapy (ACT) with tumor-infiltrating lymphocytes (TILs) was recently approved by the United States Food and Drug Administration (FDA) for the treatment of cutaneous metastatic melanoma resistant to immune checkpoint inhibitor (ICI) therapy(1). In parallel, the first randomized phase 3 trial of TIL therapy in melanoma, demonstrated superior progression-free survival compared to ipilimumab, further supporting the clinical potential of TILs(2). Despite its proven efficacy, the number of patients eligible for TIL therapy is limited by the need for accessible tumor tissue, the risk of unsuccesfull TIL outgrowth and the highly toxic pre- and post-conditioning regimens.

Expansion and/or modification of T cells from peripheral blood has gained growing attention(3), and constitutes an attractive non-invasive alternative to tumor resection. In support, it is reported that circulating tumor neoantigen-reactive CD8+T cells can be detected in the peripheral blood of patients with metastatic cancers(4). However, circulating endogenous tumor-reactive T cells are rare. Artificial antigen-presenting cells (aAPCs) to selectively expand tumor-specific T cells represent an attractive and cost-effective strategy to expand such T cells and numerous technologies have been developed over the past decades(5,6). These include cellular aAPCs, derived from established animal or human malignant cell lines, and acellular approaches using microbeads or nanoparticles for *ex vivo* expansion(7). Their use is limited by the need to remove the allogenic component or beads post-expansion, providing additional stress and handling to the final T cell product. To overcome these challenges several biomaterial-based aAPCs have recently been developed, many of which represent complex structures and include biomaterials with no previous records of clinical use(7).

We have developed a novel, simple, and easily modifiable aAPC technology(8). This technology is developed for selective expansion of low frequent antigen-specific T cell populations directly from peripheral blood(8). Antigen-presenting scaffolds (Ag-scaffolds) comprise streptavidin-conjugated dextran backbone on which biotinylated molecules can be attached. Dextran is already widely used in clinical settings as it has the attractive feature of being biodegradable(9). Accordingly, Ag-scaffolds are degraded in the culture when used for T cell expansion, with no removal step prior to clinical use required. Peptide-major histocompatibility complex (pMHC) are attached to the Ag-scaffold together with interleukins 2 and 21 (IL-2 and IL-21), which allows for the selective stimulation of specific T cells recognizing the given pMHC(8).

In the present study, we used Ag-scaffolds to expand circulating T cells specific to a selection of 30 shared human leucocyte antigens (HLA)-A*02:01-restricted tumor associated antigens (TAAs) in melanoma. The peptides were selected as the 30 most frequently detected TAA-specific T cell populations in a cohort of HLA-A*02:01-positive melanoma patients (n = 87), in which 80% showed reactivity to at least one of the 30 selected peptides(8). The Ag-scaffold technology enables the simultaneous expansion of numerous antigen-specific T cell populations from a single sample. In a cohort of HLA-A*02:01-positive patients with metastatic melanoma, Ag-scaffolds were shown to expand TAA-specific T cells from more than 60% of the patients. The resulting multiple antigen-specific endogenously derived T cell (MASE-T) product showed reactivity when challenged with cognate antigen and killing capacity when co-cultured with established melanoma cell lines(8).

In this, first-in-human phase I clinical trial, six patients with ICI-resistant metastatic melanoma were treated with a single MASE-T infusion, following three days of pretreatment with lymphodepleting chemotherapy consisting of cyclophosphamide and fludarabine phosphate. The primary endpoint was the safety and feasibility of the treatment, while secondary endpoints included clinical efficacy, characterization of the MASE-T infusion product and the *in vivo* persistence of the MASE-T cells.

## Methods

### Pre-screening and screening procedures

Criteria for inclusion and exclusion are listed in Supplementary Paragraph S1. All referred patients were pre-screened for HLA-A*02:01-positivity by flow cytometry performed on a single blood test and only HLA-A*02:01-positive patients continued with the screening procedures. Positive results were later confirmed by PCR. HLA-A*02:01-positive patients were tested for the presence of TAA-specific CD8+T cells by incubating PBMCs with PE/APC-labeled tetramers loaded with the 30 selected melanoma TAA peptides (Supplementary Table S1) for 15 min. at 37°C in the presence of 50nM dasatinib to inhibit TCR internalization. The generation of pMHC and tetramers is described in Supplementary Paragraph S2A. Following tetramer-staining, cells were stained with live/dead marker (LIVE/DEAD Fixable Near-IR, Invitrogen 2451278) and antibodies towards the surface markers CD3 and CD8 at 4°C for 30 min (Supplementary Table S2). Only patients with an identifiable population of tetramer-positive T cells (>0.002% of CD8+cells identified by at least 10 events) were included in the trial. Finally, inclusion criteria were adjusted for the last two included patients to only include patients with a CD8+T cell frequency >10%.

### MASE-T expansion

The assembly of the Ag-scaffolds is illustrated in Figure 1A and described in Supplementary Paragraph S2B. The Ag-scaffolds underwent further identity testing as detailed in Supplementary Paragraph S2C.

**Figure 1.**
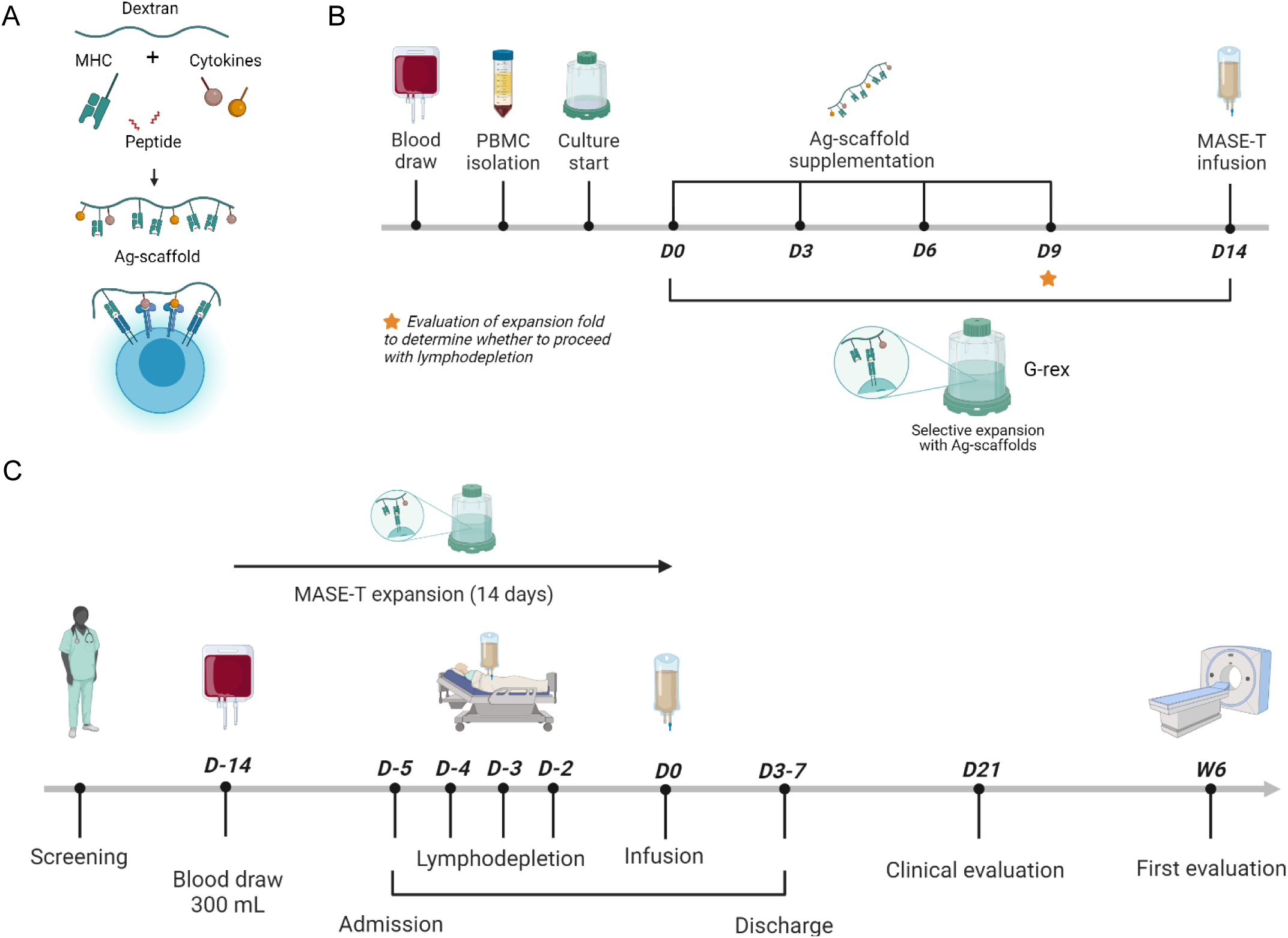
Ag-scaffold-expansion of multiple antigen-specific endogenous-derived T cells (MASE-T) for adoptive transfer in patients with metastatic melanoma. **(A)** Schematic drawing of an antigen-presenting scaffold (Ag-scaffold) engaging with an antigen-specific T cell via specific peptide-MHC complexes (pMHCs) attached to a dextran backbone, along with co-attached cytokines IL-2 and IL-21 for optimal T cell stimulation. **(B)** MASE-T expansion schedule. On day 0, 300mL of blood is drawn from the patient, and PBMCs isolated and cultured in Grex containers in the presence of Ag-scaffolds. An expansion fold ≥5 on day 9 allows for initiation of lymphodepleting chemotherapy and subsequent MASE-T cell infusion of day 14. **(C)** Trial design. Patients included after the initial screening have blood drawn for MASE-T expansion on day -14, and is admitted for lymphodepleting chemotherapy five days prior to MASE-T administration on day 0, if the fold-expansion evaluation is passed. After MASE-T treatment, patients are followed up for clinical and response evaluation.

For MASE-T production peripheral blood mononuclear cells (PBMCs) were isolated from 300 mL peripheral blood using Lymphoprep (Stemcell Technologies®) density gradient centrifugation on day 0. PBMCs were transferred to a G-Rex 100system (Wilson-Wolf) and grown in X-VIVO 15 media (Lonza) containing 5% heat-inactivated human AB serum (Sigma), antibiotics (Gentamicin/Amphotericin, Thermo Fisher Scientific) and Ag-scaffolds (0.16 nM prefiltration). During the 14-day expansion period, the cell culture was supplemented with fresh media and Ag-scaffolds on day 3, 6, and 9.

The MASE-T culture was assessed for the presence of TAA-specific CD8+T cells by tetramer-staining on day 9. An expansion fold of >5 was required for the patients to proceed with hospital admission and initiation of lymphodepleting chemotherapy, whereas upon an expansion fold ≤5, the expansion was extended with one additional stimulation on day 12 and infusion on day 17 (given an expansion fold >5 on day 12) (Figure 1B). On day 14, cells were harvested, resuspended in NaCl and 2.5% human serum albumin (CLS, Bering) and immediately administered to the patient (Figure 1B and 1C).

### Treatment Schedule

The treatment schedule is outlined in Figure 1C. Included patients were subjected to a blood draw (300 mL) performed in an outpatient setting in a 450 mL blood bag (Fresenius Kabi). The blood was transported directly to the clean room facility for immediate isolation of PBMCs and subsequent stimulation with Ag-scaffolds. Nine days post blood draw, the patient was admitted for lymphodepleting chemotherapy with three and two administrations of cyclophosphamide (500 mg/m2, days -4, -3, and -2) and fludarabine phosphate (30 mg/m^2^, days -4 and -3), respectively. The final MASE-T product was infused on day 0 followed by administration of pegfilgrastim (6 mg). The patients did not receive postconditioning IL-2. Patients were discharged when deemed clinically safe by the treating physician. After discharge, patients were clinically evaluated on day 21, while the first response evaluation was performed six weeks post MASE-T therapy.

The primary endpoints were to assess the safety and feasibility of the treatment evaluated by Common Terminology Criteria for Adverse Events (CTCAE) version 5.0(10). Secondary endpoints included characterization of the T cell profile, persistence of the infused MASE-T cells *in vivo,* and clinical efficacy evaluated as Best Overall Response (BOR) according to RECIST 1.1, Progression Free Survival (PFS) and Overall Survival (OS).

The trial was scheduled to include 12 patients. However, it was decided to terminate it prematurely for further cell product optimization.

### Phenotyping of MASE-T products and PBMCs

Phenotypic characterization of the MASE-T infusion products and PBMCs is described in Supplementary Paragraph S3A.

### Intracellular cytokine staining

Tumor reactivity of MASE-T infusion products and YT was assessed by intracellular cytokine staining as described in Supplementary Paragraph S3B.

### Data collection and data analyses

Patient enrollment and treatment as well as data collection was conducted at the National Center for Cancer Immune Therapy, Department of Oncology, Herlev Hospital, Denmark, Study data were collected and managed using Research Electronic Data Capture (REDCap) electronic data capture tools hosted at Region Hovedstaden(11,12).

Analyses of clinical data were performed in R Studio (version 4.3.2). The packages used for data analyses were; ggplot2(13), ggsurvfit(14), tidiverse(15) and gt(16). Translational data was evaluated using GraphPad version 10. The flow cytometry data, including Umap and FlowSOM analysis, was performed using FlowJo™ version 10 software (BD Life sciences)(17–19). *Statistics*: Data cut-off was April 25, 2025. Survival curves were calculated using the Kaplan-Meier method. Correlation analysis for the comparison of characteristics of MASE-T products belonging to the two RECIST categories, progressive disease (PD) and stable disease (SD) were performed using two-sample T test.

## Results

### Generation of MASE-T products for patients included in the trial

The Ag-scaffold pool used for the generation of the MASE-T products was validated and stability tested as described in Supplementaray Paragraph S4 and illustrated in Supplementary Figures S1A-C and S2A-C.

A total of 21 patients with stage IV, ICI-resistant metastatic melanoma entered the trial screening procedures between August 2021 and March 2024 (Figure 2A). Of these, 13 patients were HLA-A*02:01-positive based on antibody-based HLA-typing (Supplementary Figure S2D) and twelve had an identifiable baseline population of TAA-specific T cells, making them eligible for inclusion (Supplementary Figure S2E). However, four of the twelve patients were excluded before MASE-T production due to CNS metastases (MM2011.03), rapid cancer progression (MM2011.08 and 17), or difficulties with the blood draw (MM2011.07). One patient was excluded after MASE-T production due to a delayed response to prior treatment (MM2011.13) (Figure 2A). The first included patient (MM2011.02), was excluded due to challenges with the culture conditions, and despite extending the expansion period, the expansion criterion was not met on day 12. This prompted an evaluation and optimization of the expansion procedure, including an increase in culture vessel capacity. The revised setup supported higher cell numbers at culture initiation and resulted in improved expansion of virus-specific T cells from healthy donor PBMCs (Supplementary Figure S2F). Following optimization, patient MM2011.02 was re-included (MM2011.04), resulting in successful infusion of MASE-T cells (Figure 2A).

**Figure 2.**
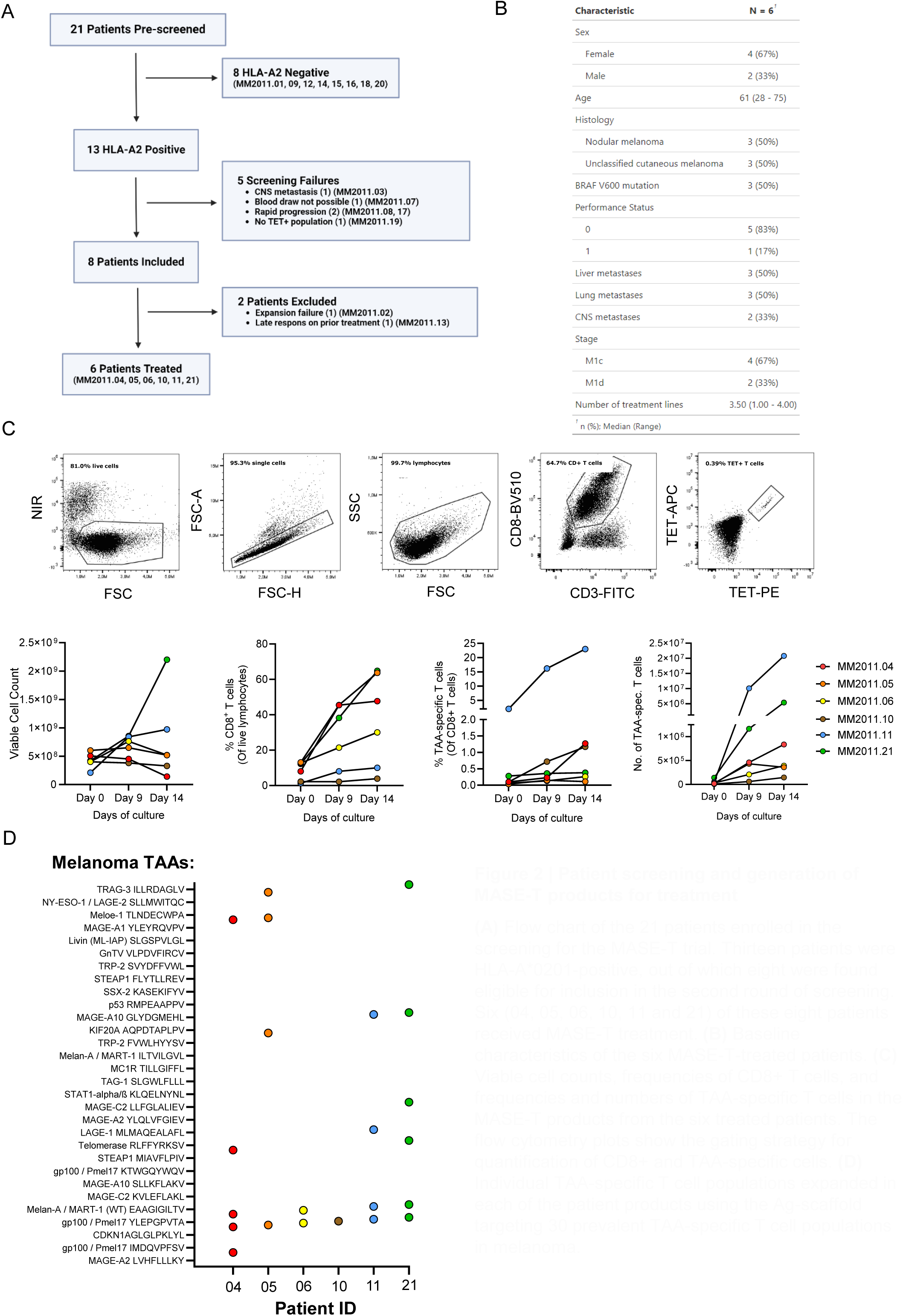
Patient screening and generation of MASE-T products for treatment. **(A)** Flow chart of the 21 patients enrolled in the screening for the MASE-T trial. Thirteen patients were HLA-A*0201-positive, out of which eight were found eligible for inclusion in the second round of screening. Six (04, 05, 06, 10, 11 and 21) of these eight patients received MASE-T treatment. **(B)** Baseline characteristics of the six MASE-T-treated patients. **(C)** Viable cell counts, frequencies of CD8+ T cells, and frequencies and numbers of TAA-specific T cells in the MASE-T products from the six treated patients. The flow cytometry plots show the gating strategy for quantification of CD8+ and TAA-specific cells. **(D)** Individual TAA-specific T cell populations expanded in each of the patient products using the Ag-scaffold targeting 30 prevalent TAA-specific T cell populations in melanoma.

Thus, MASE-T expansion was feasible in 88% (7/8) of the patients in the intention-to-treat group, and after optimization, MASE-T expansion was feasible in all the patients (7/7), of which six received the treatment. All six MASE-T products passed the expansion criteria on day 9 (fold expansion >5 of TAA-specific CD8 T cells) and all patients received the product after 14 days of expansion (Supplementary Figure S3A).

Baseline characteristics of the six treated patients are shown in Figure 2B. The median age was 61 years (range: 28-75 years). Four patients had stage M1c disease with organ metastases (liver, lung, intestine, or adrenal glands), and two patients had stage M1d disease (brain metastases). Individual patient characteristics are shown in Supplementary Figure S3B. Overall, patients were heavily pretreated with a median of 3.5 prior treatment lines (range: 1-4). All patients had PD1-inhibitor resistant disease and 5/6 had previously received ipilimumab. Three patients had BRAF V600-mutated tumors and had progressed on BRAF/MEK inhibitor therapy. Further, two patients (MM2011.04 and MM2011.10) had received Temozolomide before inclusion in the trial. Notably, the frequency of CD8+T cells decreased markedly between the initial inclusion of MM2011.02 and expansion day 0 (Day -14) of MM2011.04 (i.e, the re-included patient MM2011.02), suggesting a lymphodepleting effect of Temozolomide (Supplementary Figure S3C). Supporting this observation, patient MM2011.10 also exhibited a relatively low frequency of CD8+T cells at inclusion (Supplementary Figure S3D), which partially recovered in both patient MM2011.04 and patient MM2011.10 prior to MASE-T infusion (Supplementary Figure S3E).

The MASE-T products from the six treated patients were quantified and tested for CD8+and TAA-specific T cells on days 0, 9 and 14. As expected, Ag-scaffold-expansion led to the enrichment of both CD8+and TAA-specific T cells in all products over the 14-day expansion period, with patients MM2011.11 and MM2011.21 exhibiting the most pronounced expansion of TAA-specific T cells (Figure 2C). The number of PBMCs added to the cultures ranged from 200-600*10^6^ and this number either decreased or remained stable in most cultures, indicating limited expansion of irrelevant T cells (Figure 2C). Patients received a median of 615.5*10^3^ (range: 146.1-20790*10^3^) TAA-specific CD8+T cells, corresponding to a median of 9.1*10^3^ (range: 1.6-219.5*10^3^) TAA-specific CD8+T cells per kg body weight (data not shown). To identify the expanded TAA-specific T cell populations we analyzed the MASE-T products using combinatorial-encoded tetramer staining (Supplementary Figure S1B). In most patients (5/6), multiple TAA-specific T cell populations were detectable in the final product with a median of four populations per patient (range: 1-6) (Figure 2D).

### MASE-T treatment tolerability and clinical efficacy

Overall, MASE-T treatment was well-tolerated with no registered serious adverse events (SAEs) and no suspected expected serious adverse reactions (SUSARs). Further, no AEs occurred in relation to the MASE-T infusion (Figure 3A). All patients experienced AEs related to the chemotherapy, with all developing grade 4 lymphopenia. Half of the patients developed neutropenia, including three cases of grade ≥3 neutropenia. Additionally, one patient experienced grade 3 anemia. The median admission time was nine days (range: 8-13).

**Figure 3.**
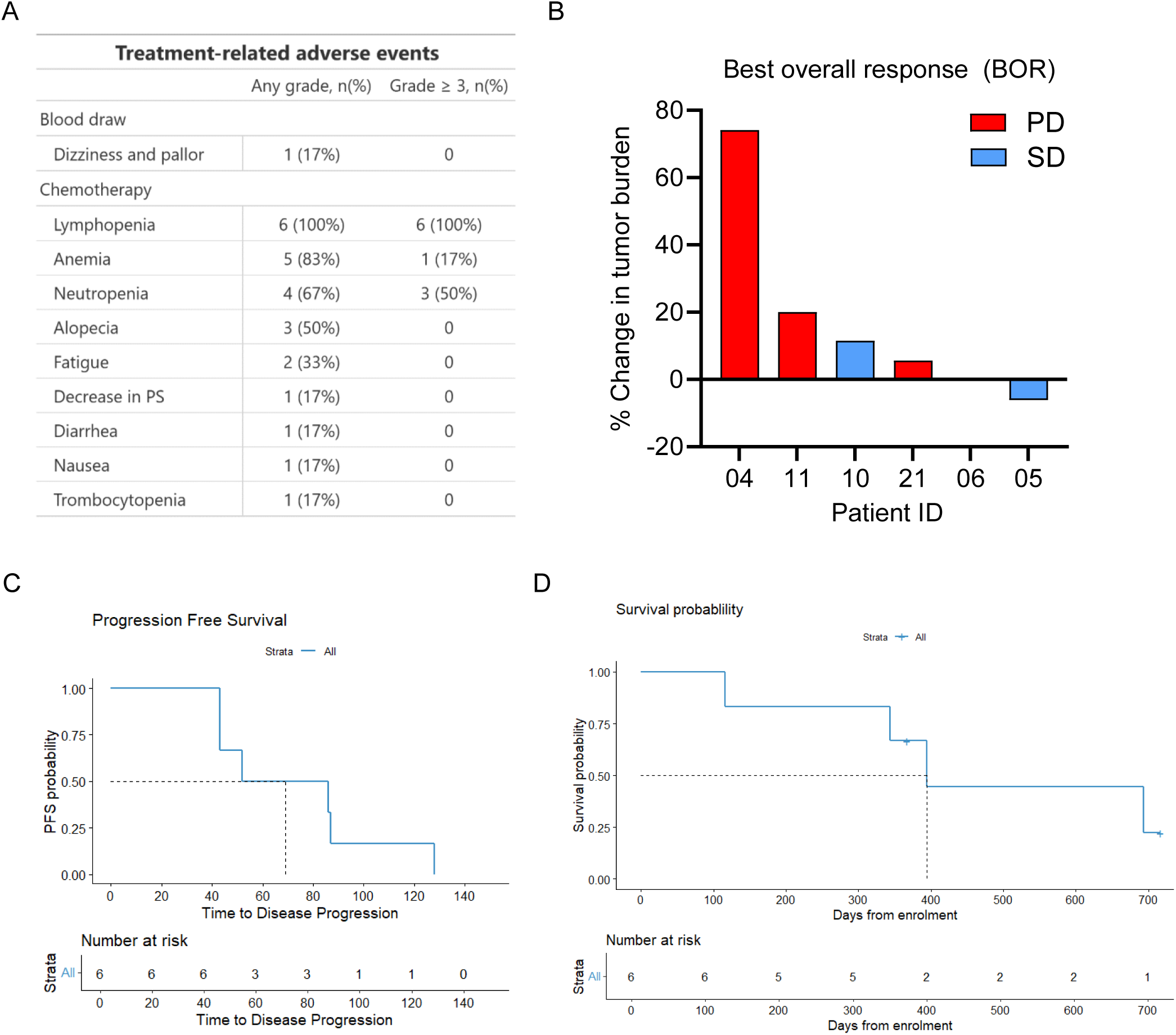
Treatment-related adverse events (AEs) and clinical response in MASE-T-treated patients. **(A)** Treatment-related AEs. No AEs related to the MASE-T infusion were registered. Most AEs were related to the chemotherapy. **(B)** Best overall response (BOR) evaluated by RECIST 1.1, PD: Progressive disease, SD: Stable disease. Three patients had SD as BOR, but for only one patient SD was confirmed on the second scan 3 months post MASE-T treatment. **(C)** Median progression-free survival (PFS) was 69 days and median overall survival (OS) **(D)** was 394 days in MASE-T-treated patients

All six MASE-T-treated patients were evaluable for clinical response; three had PD on the first scan performed six weeks after MASE-T infusion, while the remaining three patients had SD as BOR (Figure 3B). In one patient (MM2011.06), SD was confirmed on the second scan three months post-treatment, but the patient developed PD after 4.2 months. Only one patient (MM2011.05) demonstrated a minor decrease in tumor burden (6%). Accordingly, the median PFS was 69 days, while median OS was 394 days (Figure 3C-D). Five of the six MASE-T-treated patients received subsequent therapy following MASE-T treatment (Supplementary Figure S3F). Notably, two patients (MM2011.06 and MM2011.11) underwent TIL therapy with lymphodepletion and post-conditioning IL-2 after MASE-T, with one patient (MM2011.11) achieving a partial response (PR). The remaining patients did not respond to any treatment administered after MASE-T therapy, and at data cut-off, four of the six patients had deceased.

### MASE-T product characteristics and correlation to best overall response

Due to the limited number of patients, comparison of MASE-T product characteristics between the two RECIST categories, PD and SD (Figure 4A-B and Supplementary Figure S4A-B) should be interpreted with caution. Interestingly, MASE-T products from patients with PD tended to exhibit higher viable cell counts, greater total numbers of TAA-specific T cells, and a broader range of TAA specificities compared to those from patients with stable disease (Figure 4A). In contrast, products from patients with SD did exhibit a lower frequency and reduced expression of PD-1 among expanded TAA-specific T cells compared to those from patients with PD (Figure 4B). Expression levels of CD28, CD39, TCF-1, KI67, and GZMB, assessed by both mean fluorescence intensity (MFI) and the frequency of marker-positive TAA-specific cells, indicated an active cytotoxic and proliferative profile. However, no significant differences were observed between patients with PD and those with SD (Supplementary Figure S4A-B).

**Figure 4.**
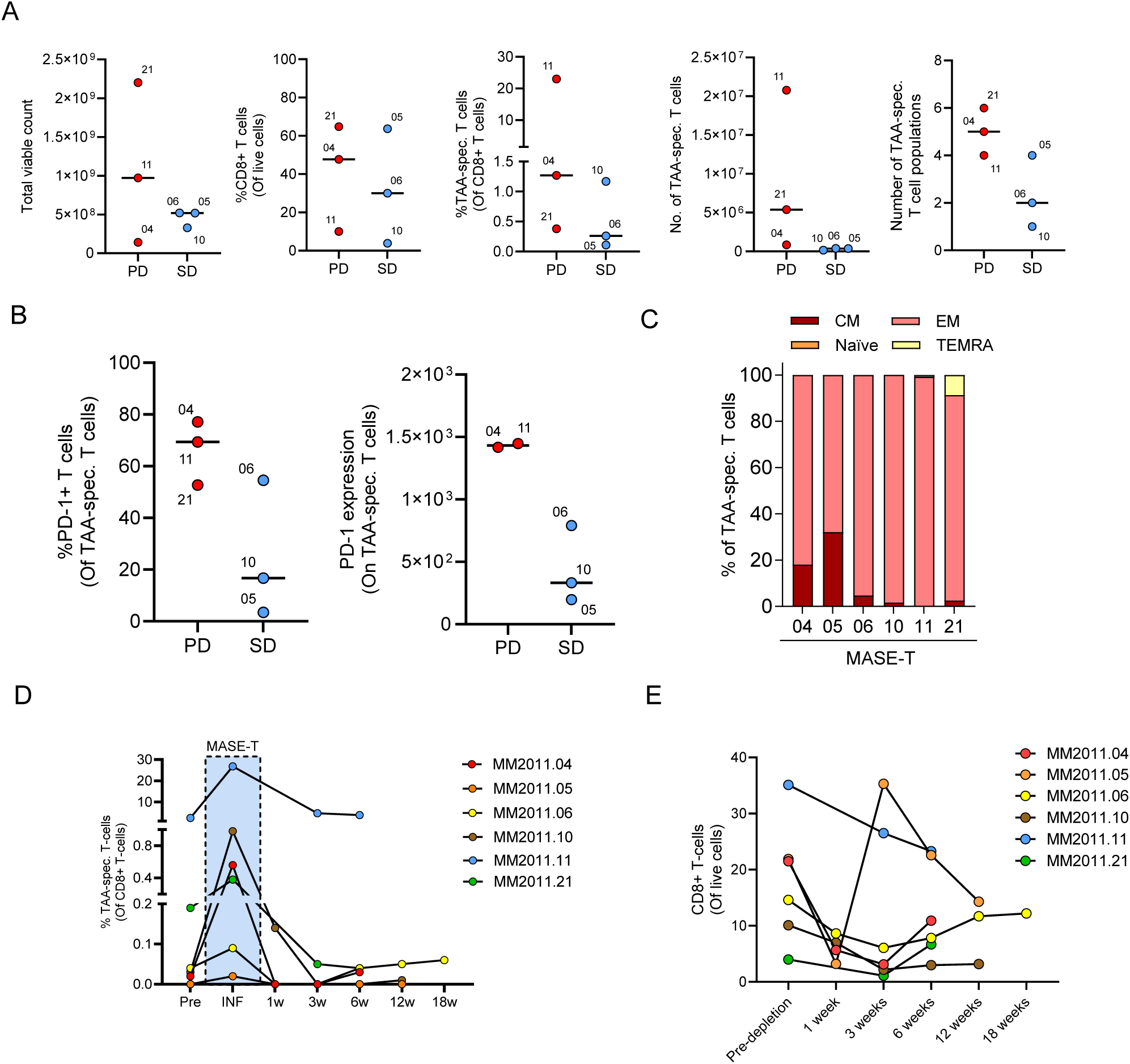
MASE-T product characteristics and correlation to best overall response. **(A)** Total viable cell counts, frequencies of CD8+ T cells, frequencies and numbers of TAA-specific cells, and numbers of TAA-specific T cell populations following grouping of MASE-T products according to the RECIST 1.1 classification of the patients. Horizontal lines represent the mean number/frequency. **(B)** Frequency and mean fluorescence intensity (MFI) of PD-1 expression on TAA-specific cells in MASE-T products grouped according to the RECIST 1.1 classification of the patients. Horizontal lines represent the mean frequency/intensity. **(C)** Subdivision of TAA-specific cells in the MASE-T cell products into naïve-like (Naïve), central memory (CM), effector memory (EM) and effector memory that re-express CD45RA (TEMRA) based on expression of CD45RA and CCR7. **(D)** Frequency of TAA-specific cells in the infusion product (INF), and in peripheral blood prior to (Pre) and at various time points after (1w, 3w, 6w, 12w and 16w) MASE-T treatment. **(E)** frequencies of CD8+ T cells in PBMCs of MASE-T treated patients pre-lymphodepletion and at various time points after MASE-T treatment.

Based on CD45RA-CCR7 characterization, most expanded TAA-specific T cells were of the effector memory (EM) T cell subtype in all MASE-T products (Figure 4C).

To assess the persistence of the infused MASE-T products, the frequency of TAA-specific T cells in peripheral blood (Figure 4D) was measured. Analyses at one-and three-weeks post-ACT were compromised by lymphodepletion, which led to a marked reduction in circulating lymphocytes (Supplementary Figure S4C). Although the lymphocyte population, and in particular CD8+T cells, gradually recovered over time, levels did not return to those observed prior to lymphodepletion (Figure 4E and Supplementary Figure S4D). Despite the effect of lymphodepletion, TAA-specific T cells remained detectable in the peripheral blood of five out of six patients at six weeks post MASE-T infusion (Figure 4D). In two patients (MM2011.06 and MM2011.11), the frequency of TAA-specific T cells was sufficient to permit phenotypic analysis. These analyses revealed that the expanded TAA-specific T cells within the MASE-T product displayed a distinct phenotypic profile compared to those in peripheral blood, and appeared to gradually revert toward their pre-expansion phenotype over the course of 3-18 weeks post infusion (Supplementary Figure S4D).

### Comparison of MASE-T and TIL products from patients receiving TIL therapy following MASE-T treatment

Two patients (MM2011.06 and MM2011.11) received TIL therapy following MASE-T treatment. At cancer progression 18 weeks after MASE-T infusion, patient MM2011.06 was re-treated with a BRAF/MEK inhibitor but experienced progression again after eight months. Subsequently, the patient was referred to TIL therapy and received TIL therapy with 55.7×10^9^ cells 19 months post MASE-T infusion (Supplementary Figure S3F). Unfortunately, the patient progressed shortly after TIL infusion and died 23 months post MASE-T therapy. In contrast, patient MM2011.11 progressed immediately after MASE-T therapy and was referred for TIL treatment, with tumor resection performed five months after MASE-T therapy. TIL treatment with 36.9×10^9^ cells was administered eight months post MASE-T resulting in a PR that lasted for more than one year (Supplementary Figure S3F).

For these two patients, we performed phenotypic profiling of the MASE-T infusion product, the expanded TIL (REP-TIL) infusion product and, for patient MM2011.11, also the young TILs (yTIL) cultured directly from a tumor biopsy obtained prior to MASE-T therapy. All products consisted mainly of CD3^+^ T cells (Supplementary Figure S5A), however, the distribution of CD4+and CD8+T cells varied. In patient MM2011.06, the REP-TIL product consisted almost exclusively of CD4+T cells, whereas the MASE-T product was mainly CD8+T cells (Figure 5A). In patient MM2011.11, the yTIL from the pre-MASE-T biopsy were predominantly CD8+T cells, whereas the MASE-T and the REP-TIL infusion product contained 40% and 20% CD4+T cells, respectively (Figure 5B). Among the CD4+T cells, the REP-TIL products exhibited a higher frequency of FoxP3^+^ regulatory T cells (Tregs) compared to their corresponding MASE-T products, which was further supported by increased expression of the proliferation marker ki67 on the regulatory T cell (Treg) population (Supplementary Figure S5B). This may reflect the influence of IL-21 during MASE-T expansion, as IL-21 has been shown to suppress IL-2-induced Treg expansion(20).

**Figure 5.**
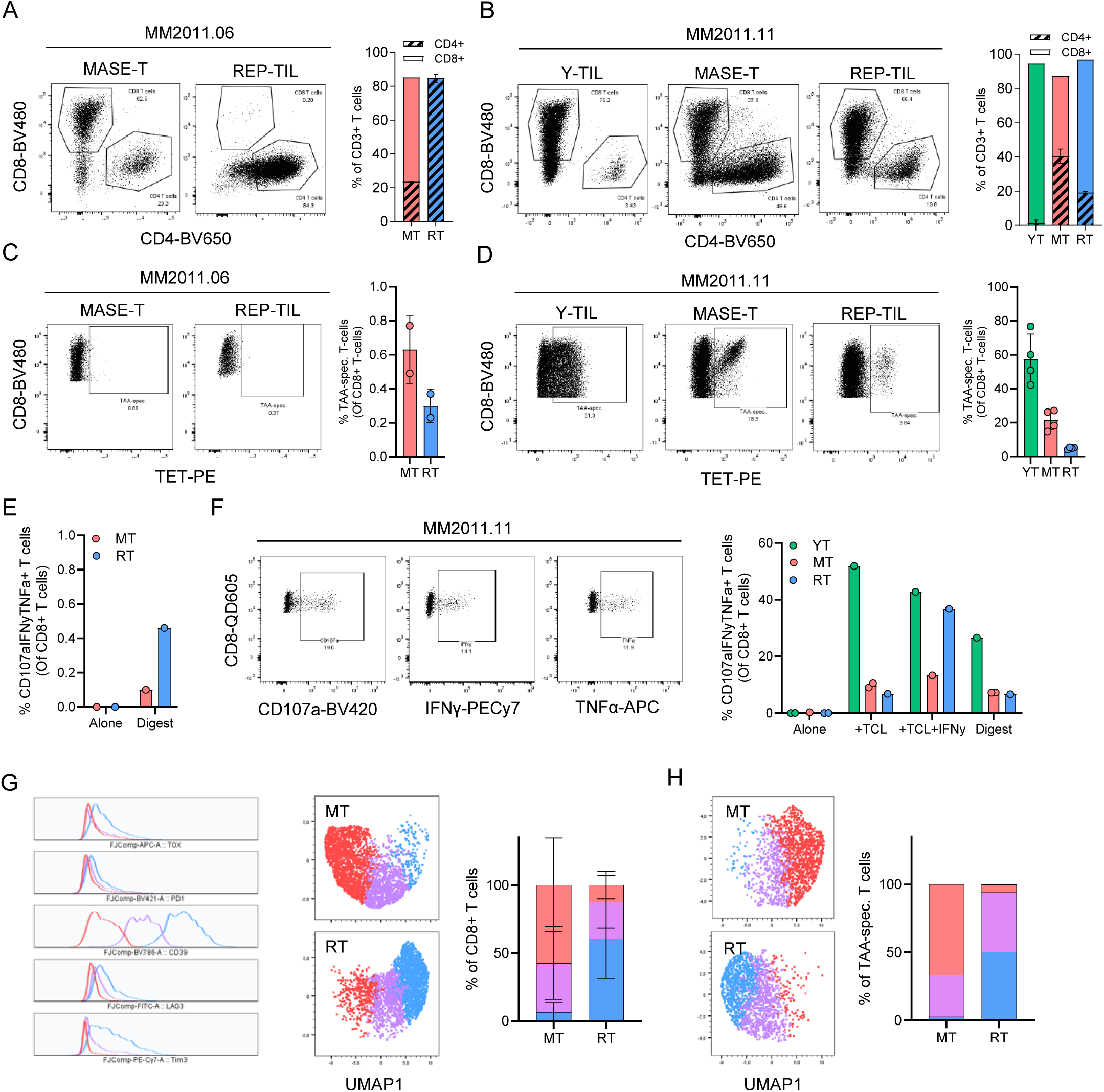
Comparing the MASE-T and TIL products in patients that received both. **(A and B)** Frequency of CD4^+^ and CD8^+^ T cells in Y-TIL (YT) from a pre-MASE-T tumor biopsy, MASE-T (MT) infusion product and REP-TIL (RT) infusion products from patients **(A)** MM2011.06, and **(B)** MM2011.11. Bar graphs display the mean +/-SD frequency of CD8 and CD4 cells out of total CD3+ T cells. **(C and D)** Frequency of TAA-specific T cells in YT, MT, and RT from patients **(C)** MM2011.06, and **(D)** MM2011.11. Bar graphs display the mean +/- SD frequency of TAA-specific cells out of CD8+ T cells. **(E and F)** Frequency of cytokine-producing (TNF and IFNg) and CD107a-expressing CD8+ T cells in YT, MT and RT from patients **(E)** MM2011.06, and **(F)** MM2011.11 following in vitro-restimulation with autologous tumor cell lines with or without prestimulation with IFNg (TCL and TCL+IFNg, respectively), or tumor digest (Digest), or without re-stimulation (Alone). Bar graphs display the frequency of multi-functional (CD107a+ IFNg+ TNF+) cells. **(G and H)** FlowSOM analysis performed on **(G)** CD8+ T cells in MT and RT from patient 06 and 11 (pooled), and **(H)** TAA-specific T cells in MT and RT from patient 11.

As previously shown, only a limited fraction of the CD8+T cells in both the MASE-T and TIL products from patient MM2011.06 were specific for the Ag-scaffold-targeted TAA epitopes (Figure 5C). In contrast, the MASE-T product from patient MM2011.11 contained a substantial population of TAA-specific CD8⁺ T cells (∼20%), predominantly targeting MART-1 (Figure 5D and Figure 2C-D). Interestingly, this patient also exhibited a ∼5% TAA-specific T cell population in the REP-TIL product, which may have been influenced by the preceding MASE-T infusion (Figure 5D). However, MART-1-specific T cells were also detected in TILs expanded from a biopsy obtained prior to MASE-T therapy (Figure 5D and Supplementary Figure S5C). Furthermore, a circulating TAA-specific T cell population of approximately 2% was present at inclusion, indicating that the patient had both circulating and tumor-infiltrating MART-1-specific T cells prior to receiving MASE-T treatment (Supplementary Figure S2E).

The functional capacity of the products from these two patients was evaluated. For patient MM2011.06, only tumor digest was available for functional assessment due to unsuccessful establishment of a TCL. In co-culture assays, both the MASE-T and REP-TIL products exhibited low reactivity, as indicated by the limited frequency of T cells co-expressing both TNF, IFNγ and CD107a in response to the tumor digest (Figure 5E). The slightly higher reactivity in the REP-TIL product may be mediated by other tumor specificities than those targeted by the Ag-scaffold (Figure 5E). In general, a higher frequency of tumor-reactive T cells was observed in the yTIL, MASE-T and REP-TIL product from patient MM2011.11 upon co-culture with TCL and digest, reflecting the frequency of TAA-specific T cells found in the products (Figure 5D and 5F). However, co-culture with a TCL preincubated with IFNγ markedly enhanced the reactivity of the REP-TIL product, suggesting that IFNγ may induce the presentation of alternative antigens on the TCL that were specifically recognized by the REP-TIL product (Figure 5F). Additionally, the highest reactivity was observed in the yTILs, independent of IFNγ-stimulation, suggesting that these recognises a more diverse antigen repertoire.

Finally, a phenotypic characterization of the MASE-T and TIL products was performed on total CD8+T cells from patients MM2011.06 and MM2011.11, as well as on TAA-specific T cells from patient MM2011.11. FlowSOM analysis revealed that both total CD8+and TAA-specific T cells in the MASE-T and REP-TIL products clustered into distinct phenotypic groups (Figure 5G and H). CD8+T cells and TAA-specific T cells in the MASE-T products were predominantly associated with a cluster characterized by low expression of exhaustion markers including CD39, Tox, PD1, LAG3 and Tim-3. In contrast, the dominating cluster in the REP-TIL products exhibited higher expression of these markers (Figure 5G and H and Supplementary Figure S5D).

## Discussion

The FDA approval of TIL therapy for metastatic melanoma marks an essential step in the development of cellular therapies for solid tumors. However, while TIL treatment can be highly effective and provide long-lasting responses, most patients are still not eligible or fail to respond to therapy(2,21). The need for resectable tumor tissue constitutes a significant limitation, particularly in cancers that seldom metastasize to skin or lymph nodes. Even when resection is feasible, unsuccessful TIL outgrowth from tumor biopsies remains a frequent challenge, preventing product generation in a subset of patients. Furthermore, the process of tumor resection, TIL expansion, and subsequent infusion is logistically complex and time-consuming, often requiring several weeks - a major issue for patients with rapidly progressing disease. Thus, there is a need for alternative expansion strategies to increase the accessibility and efficacy of ACT in solid tumors.

From a logistical perspective, MASE-T therapy represents an attractive alternative to traditional TIL therapy. Blood draw is possible from almost any patient and can be performed without laborious planning. Furthermore, it is a minimally invasive procedure with a low complication rate(22). Using the Ag-scaffold technology, expansion of TAA-specific T cells from peripheral blood was feasible in all included patients, providing a T cell product containing multiple expanded endogenous TAA-specific T cell populations. Even in patients with a limited frequency of TAA-specific T cells at baseline we achieved an expansion fold of at least six on expansion day 14, demonstrating the potential of the Ag-scaffold technology to capture also very low frequent cell populations.

Infusion of the MASE-T product did not give rise to any AEs, indicating that the treatment was well tolerated and safe. Toxicities were almost exclusively related to the chemotherapeutical regimen used for lymphodepletion, which was achieved in all patients,. All AEs related to the chemotherapy were manageable and most were mild. Thus, compared to conventional TIL therapy, MASE-T treatment has an attractive toxicity profile, likely due to the dose-reduced chemotherapy and lack of IL-2 post-conditioning. While preconditioning lymphodepletion serves several purposes, including eradication of Tregs, removal of cellular cytokine sinks(23), and to create physical space for the infused TIL product(24), post-conditioning IL-2 administration enhances the anti-tumor response by increasing *in vivo* persistence and activation of the infused cells(25). Thus, the reduced pre- and post-conditioning regime may reduce the persistence of the infused MASE-T product and could potentially limit the clinical response.

In addition to the potential impact of the reduced pre- and post-conditioning regime, several additional factors may have contributed to the limited clinical efficacy of MASE-T therapy. The patient population was characterized by advanced-disease stages (M1c-d), high tumor burden, and a high number of prior treatment lines (median: 3.5), all of which are factors known to negatively impact response to TIL therapyare(26–28).

Two patients (MM2011.04 and MM2011.10) received Temozolomide shortly before MASE-T treatment. While Temozolomide is known to induce lymphopenia, its specific impact on the CD8+T cell compartment remains unclear(29). Nevertheless, both patients displayed a low proportion of CD8+T cells in peripheral blood at the time of MASE-T, suggesting that Temozolomide may reduce the availability of CD8+T cell precursors necessary for robust TAA-specific MASE-T expansion. Finally, the poor prognosis of the included patients is further illustrated by the fact that five out of six patients did not respond to any other therapy administered post MASE-T therapy.

The absolute number of infused MASE-T cells (median 504×10^6^, range: 130-2200×10^6^) was significantly lower than the number typically reported in TIL therapy (range: 1-110×10^9^)(2,21), where a high number of infused cells and, in particular, a high number of CD8+ T cells has been correlated with a favorable response across tumor subtypes (26,30,31). In CAR-T therapy, a working dose of 20-50×10^6^ cells is often sufficient to elicit clinical responses, even in patients with a relatively high tumor burden(32). However, these dosing guidelines are primarily based on hematological malignancies, where CAR-T cells can readily access malignant cells in the circulation or bone marrow without needing to traffic to solid tumor sites or overcome an immunosuppressive tumor microenvironment. In addition to the absolute number, the number of adoptively transferred tumor-specific T cells predicts efficacy in clinical ACT studies(30,31). Despite Ag scaffold-directed enrichment for TAA-specific CD8+ T cell specificities, the MASE-T product contained a limited number of TAA-specific cells with a median of 9×10^3^ (range: 1.6-219×10^3^) TAA-specific cells/kg body weight. In comparison, the number of tumor reactive CD8+ T-cells in TIL products from responding patients often exceeds 1×10^8^ cells(30). Thus, considering the high tumor burden in the included patients, the MASE-T cells may have been too few to make a difference. However, not only the absolute cell count, but also the phenotype of the tumor-specific T cells is of importance for the outcome(33,34).

Most of infused MASE-T cells displayed an effector memory phenotype, consistent with previous observations in TIL therapy. In contrast, the REP-TIL product from patients MM2011.06 and MM2011.11 had a higher frequency of T-regs with elevated Ki67 expression, likely reflecting the extensive stimulation involved in the expansion of TILs.

Phenotypic comparison of the MASE-T and REP-TIL products from patients MM2011.06 and MM2011.11 revealed that the infused MASE-T cells (both total CD8+and TAA-specific) exhibited a less exhausted phenotype. These cells were predominantly characterized by lower expression of CD39 and other exhaustion markers compared to the majority of cells in the corresponding REP-TIL products. Despite these phenotypic differences, the *in vitro* responses of the MASE-T and REP-TIL products to stimulation with TCL without IFNγ and with tumor digest largely reflected the frequencies of TAA-specific cells in the products. This suggests that the phenotypic differences may be more relevant to other functional properties, such as *in vivo* persistence, rather than antigen-specific responsiveness.

The success of any aAPC strategy depends on the antigen(s) selected for T cell stimulation. Neoantigens are attractive because they are not expressed by healthy tissue. However, prediction and identification of immunogenic patient-specific neoantigens require access to tumor and healthy tissue, high-performing prediction algorithms, and high-throughput pMHC-driven T cells screening, which collectively is both time-consuming and expensive(35).

Targeting TAAs constitutes a simpler and cheaper alternative. TAAs are attractive targets for immunotherapies since they are shared among patients with cancer, allowing off-the-shelf therapies, rather than cumbersome personalized solutions(36).Supporting the relevance of targeting TAAs in melanoma, TCR-transduced T cells from peripheral blood targeting different TAAs, including MART-1, have demonstrated clinical efficacy in melanoma patients(37,38).

However, T cells genetically modified to express a high-affinity TAA-specific TCR can cause severe on-target-off-tumor toxicity, which has been observed in several clinical trials with different TAA targets(37,39,40). Endogenous TAA-specific T cells do not carry the same risks, since their TCRs have undergone negative selection in the thymus and do not react to the limited TAA expression in normal tissues. However, their low affinity may also explain the disappointing results of vaccine trials targeting TAAs(41). Nevertheless, endogenous TAA-specific T cells are frequently found in TIL infusion products from patients with melanoma and can mediate tumor cell killing(42–44). In the present study, patient MM2011.11 had a high frequency of endogenous MART-1-specific T cells in both peripheral blood and a tumor biopsy drawn prior to MASE-T treatment. However, the patient experienced progression at the time of inclusion, suggesting that either the tumor had downregulated MART-1 expression, or that the immunosuppressive TME prevented efficient tumor targeting. Supportively, the patient did not respond to a MASE-T product containing >20% Ag-scaffold-expanded MART-1-specific T cells.

Notably, the MASE-T product showed *in vitro* reactivity when co-cultured with an autologous TCL and tumor digest, suggesting that the tumor continued to express the MART-1 antigen. Furthermore, the reactivity towards autologous TCL and digest seems to correlate with the frequency of MART-1-specific T cells in the young TILs, MASE-T, and TIL products.

A limitation of the Ag-scaffold technology is that it requires pre-selection of the T cell specificities to target for expansion. This contrasts with TIL therapy, which exploits the tumor as a source of tumor-specific T cells. Thus, the relatively narrow repertoire of TAA-specific T cells found in the MASE-T products (range: 1-6, median = 4) and the lack of T cells specific for alternative antigen classes, such as neoantigens, may contribute to the limited clinical efficacy(31,44). The expansion and infusion of blood-derived neoantigen specific T cells were recently evaluated in a clinical trial involving a cohort of melanoma patients comparable to those in the current study(45). The autors demonstrated *in vitro* reactivity of the infused cells towards autologous tumor tissue in most patients, however, no clinical responses were observed (45). These findings underscore the complexity of manufacturing cellular products for solid tumors and indicate that a diverse repertoire of T cells, targering both TAAs and neoantigens may be essential for achieving therapeutic reponses.

The presence of a broader repertoire of tumor-specific T cells in the TIL compared to the MASE-T product may explain the substantial increase in reactivity of the TIL, but not the MASE-T, product from patient MM2011.11 following IFNγ-pre-conditioning of the autologous TCL. Supportively, patient MM2011.11 responded to TIL therapy after progressing on MASE-T therapy.

In a subset of solid cancers arising from viral infection, such as human papillomavirus (HPV)-driven cancers and Merkel cell carcinoma (MCC), integrated viral genes serve as attractive tumor antigens for Ag scaffold-expansion. Indeed, recent findings in MCC show that oncogenic T antigen-specific T cells could be expanded from the peripheral blood of MCC patients, with the resulting T cell products showing anti-tumor reactivity *in vitro*(46).

This trial demonstrates that Ag scaffold-expansion of MASE-T cells from patient peripheral blood is feasible and generates a well-tolerated infusion product enriched with tumor-specific CD8+T cells that can direct autologous tumor cell killing. Thus, ACT with Ag scaffold-expanded T cells represents a promising alternative when TIL therapy is not feasible, in particular for solid cancers with a well-defined antigenic landscape. However, the limited clinical efficacy observed in the current trial warrants continued optimization within several areas, including patient inclusion criteria, the size of the generated cell product, and pre-and post-conditioning regimes.

## Declarations

### Ethics approval and consent to participate

All patients provided oral and written informed consent in accordance with the declaration of Helsinki. Prior to the inclusion of the first patient, the trial was approved by the National Ethics Committee, the Danish Data Protection Agency, and the Danish Medicines Agency. The trial is registered at clinicaltrials.gov, no: NCT04904185.

### Consent for publication

Written and oral consent for publication was obtained from all participating patients.

### Availability of data and material

Any additional information required to reanalyze the data reported in this paper is available upon reasonable request.

### Competing interests

TJM, SAT, MSF, CV, MO, JWK, RBH, NJ, JSG, SKL and ÖM declares no competing interests. THB has received personal payment for lectures/presentation from Bristol Myers Squibb. ARC is co-founder of PokeAcell, developer of the Ag-scaffold technology and has stokcs or stock options in PokeAcell and Cymab. SRH is co-inventor of two patents related to the Ag-scaffold technology (EP 16205918.2 and 18178769.8) filled and owned by the Technical University of Denmark. IMS: Personal payments honoraria: received for lectures, presentations, speakers’ bureaus, manuscript writing, advisory board or educational events from MSD, Takeda, Sanofi Aventis, Janssen Cilag and BMS, Institutional grants and contracts: received from Evaxion Biotech, Adaptimmune, IO Biotech, Asgard Biotech, TILT Biotherapeutics and Enara Bio, Consulting fees: received from TILT Biotherapeutics, IO Biotech, Novartis and Genmab, Stocks/shares: IO Biotech, Meeting support: received from MSD, Clinical trial drugs: Received Relatlimab from BMS, DSMB participation: involved in only academic trials.

## Funding

Empowering cancer immunotherapy in Denmark. The Innovation Fund Denmark: <u>ImmPACT - Precision Activated Cell Therapy</u> (9122-00084B) and IMPACE (<u>0154-00037B</u>), and the Independent Research Fund Denmark: <u>EliteForsk-pris 2020</u> (9095-00029B).

## Author contributions

TJM: Preparation, creation and presentation of the published work, specifically writing the original draft and visualization/data presentation. Management of data and application of statistical analysis. Conducting a research and investigation process, specifically patient care and monitoring, performing experiments and data/evidence collection, SAT: Preparation, creation and presentation of the published work, specifically writing the original draft and visualization/data presentation. Management of data and application of statistical analysis. Conducting a research and investigation process, specifically performing experiments and data/evidence collection, MSF: Preparation, creation and presentation of the published work, specifically writing the original draft and visualization/data presentation. Management of data and application of statistical analysis. Conducting a research and investigation process, specifically performing experiments, or data/evidence collection, CV: Conducting a research and investigation process, specifically performing data/evidence collection and patient care and monitoring, MO: Development or design of methodology, JWK: Development or design of methodology, THB: Conducting a research and investigation process, RBH: Conducting a research and investigation process, specifically performing data/evidence collection, JSG: Conducting a research and investigation process, specifically performing the experiments, ARC: Development or design of methodology, SKL: Conducting research and investigation process, ÖM: Conducting a research and investigation process, SRH: Management and coordination responsibility for the research activity planning and execution. Development or design of methodology. Acquisition of the financial support for the project leading to this publication. Oversight and leadership responsibility for the research activity planning and execution, including mentorship external to the core team, IMS: Management and coordination responsibility for the research activity planning and execution. Development or design of methodology. Acquisition of the financial support for the project leading to this publication. Oversight and leadership responsibility for the research activity planning and execution, including mentorship external to the core team.

## Acknowledgements

We sincerely thank the laboratory technicians Marie Hansen, Yasmiin Akthar, Martin Jørgensen, Maria J. A. G. Westerdahl, Magnus L. Jørgensen and Majken Holm. Further, we thank the Department of Oncology, Herlev Hospital.

## List of abbreviations

ACT: Adoptive Cell Therapy
Ag-scaffolds: Antigen-presenting Scaffolds
APC: Antigen Presenting Cell
aAPC: Artificial Antigen Presenting Cell
CTCAE: Common Terminology Criteria for Adverse Events
DC: Dendritic Cell
FDA: United States Food and Drug Administration
HLA: Human Leucocyte Antigens
ICI: Immune Checkpoint Inhibitors
IFNγ: Interferon Gamma
IL-2: Interleukin 2
IL-21: Interleukin 21
MASE-T: Multiple antigen-specific endogenously derived T cells
MHC: Major Histocompatibility Complex
PD: Progressive Disease
PR: Partial Response
pMHC: Peptide-Major Histocompatibility Complex
RECIST: Response Evalution Criteria in Solid Tumors
REP: Rapid Expansion Protocol
SD: Stable Disease
TAA: Tumor-associated antigen
TCR: T cell receptor
TIL: Tumor Infiltrating Lymphocytes
TNF: Tumor necrosis Factor
Treg: Regulatory T cell

## Notes

### Competing Interest Statement

TJM, SAT, MSF, CV, MO, JWK, RBH, NJ, JSG, SKL and OM declares no competing interests. THB has received personal payment for lectures/presentation from Bristol Myers Squibb. ARC is co-founder of PokeAcell, developer of the Ag-scaffold technology and has stokcs or stock options in PokeAcell and Cymab. SRH is co-inventor of two patents related to the Ag-scaffold technology (EP 16205918.2 and 18178769.8) filled and owned by the Technical University of Denmark. IMS: Personal payments honoraria: received for lectures, presentations, speakers bureaus, manuscript writing, advisory board or educational events from MSD, Takeda, Sanofi Aventis, Janssen Cilag and BMS, Institutional grants and contracts: received from Evaxion Biotech, Adaptimmune, IO Biotech, Asgard Biotech, TILT Biotherapeutics and Enara Bio, Consulting fees: received from TILT Biotherapeutics, IO Biotech, Novartis and Genmab, Stocks/shares: IO Biotech, Meeting support: received from MSD, Clinical trial drugs: Received Relatlimab from BMS, DSMB participation: involved in only academic trials.

### Clinical Trial

NCT04904185

### Funding Statement

Empowering cancer immunotherapy in Denmark. The Innovation Fund Denmark: ImmPACT - Precision Activated Cell Therapy (9122-00084B) and IMPACE (0154-00037B), and the Independent Research Fund Denmark: EliteForsk-pris 2020 (9095-00029B).

### Author Declarations

The Danish National Committee on Health Research Ethics, 2101511

## References

1. FDA. https://www.fda.gov/news-events/press-announcement. 2024. FDA Approves First Cellular Therapy to Treat Patients with Unresectable or Metastatic Melanoma.

2. Rohaan MW, Borch TH, Kessels R, Foppen MHG, Nuijen B, Nijenhuis C, et al. Tumor-Infiltrating Lymphocyte Therapy or Ipilimumab in Advanced Melanoma. New England Journal of Medicine. 2022 Dec 8;387(23):2113–25.

3. Wang Z, Ahmed S, Labib M, Wang H, Wu L, Bavaghar-Zaeimi F, et al. Isolation of tumour-reactive lymphocytes from peripheral blood via microfluidic immunomagnetic cell sorting. Nat Biomed Eng. 2023;7(9):1188–203.

4. Yossef R, Krishna S, Sindiri S, Lowery FJ, Copeland AR, Gartner JJ, et al. Phenotypic signatures of circulating neoantigen-reactive CD8+ T cells in patients with metastatic cancers. Cancer Cell [Internet]. 2023;41(12):2154–2165.e5. Available from: 10.1016/j.ccell.2023.11.005

5. Hasan A, Selvakumar A, O’Reilly R. Artificial Antigen Presenting Cells: An Off the Shelf Approach for Generation of Desirable T-Cell Populations for Broad Application of Adoptive Immunotherapy. Adv Genet Eng. 2015;04(03):1–22.

6. Neal LR, Bailey SR, Wyatt MM, Bowers JS, Majchrzak K, Nelson MH, et al. The Basics of Artificial Antigen Presenting Cells in T Cell-Based Cancer Immunotherapies. J Immunol Res Ther. 2017;2(1):68–79.

7. Isser A, Livingston NK, Schneck JP. Biomaterials to enhance antigen-specific T cell expansion for cancer immunotherapy. Biomaterials. 2021;268.

8. Tvingsholm SA, Frej MS, Rafa VM, Hansen UK, Ormhøj M, Tyron A, et al. TCR-engaging scaffolds selectively expand antigen-specific T-cells with a favorable phenotype for adoptive cell therapy. J Immunother Cancer. 2023;11(8):1–16.

9. Dubniks M, Persson J GPO. Comparison of the plasma volume-expanding effects of 6% dextran 70, 5% albumin, and 6% HES 130/0.4 after hemorrhage in the guinea pig. J Trauma. 2009;67:1200–4.

10. U.S. Department of health and human services, National Institutes of Heallth NCI. Common Terminology Criteria for Adverse Events (CTCAE) Version 5.0 [Internet]. 2017. Available from: https://ctep.cancer.gov/protocoldevelopment/electronic_applications/ctc.htm#ctc_50

11. PA Harris, R Taylor, BL Minor, V Elliott, M Fernandez, L O’Neal, L McLeod, G Delacqua, F Delacqua, J Kirby SD. The REDCap consortium: Building an international community of software partners,. J Biomed Inform. 2009;

12. PA Harris, R Taylor, R Thielke, J Payne, N Gonzalez JGC. Research electronic data capture (REDCap) – A metadata-driven methodology and workflow process for providing translational research informatics support. J Biomed Inform. 2009;42(2):377–81.

13. Wickham H. ggplot2: Elegant Graphics for Data Analysis. Springer-Verlag New York.; 2016.

14. Terry M. Therneau PMG. GGsurvfit - Modeling Survival Data: Extending the Cox Model. Springer, New York.; 2000.

15. Wickham H, Averick M, Bryan J, Chang W, McGowan L, François R, et al. Welcome to the tidyverse. J Open Source Softw. 2019;4(43):1686.

16. Iannone R, Cheng J, Schloerke B, Hughes E, Lauer A, Seo J, et al. gt: Easily Create Presentation-Ready Display Tables. 2024.

17. McInnes L, Healy J, Melville J. UMAP: Uniform Manifold Approximation and Projection for Dimension Reduction. 2018 Feb 9; Available from: http://arxiv.org/abs/1802.03426

18. Van Gassen S, Callebaut B, Van Helden MJ, Lambrecht BN, Demeester P, Dhaene T, et al. FlowSOM: Using self-organizing maps for visualization and interpretation of cytometry data. Cytometry Part A. 2015 Jul 1;87(7):636–45.

19. Ashland, OR: Becton D and C. FlowJo^TM^ Software [software application] Version 10. 2023.

20. Kim-Schulze S, Kim HS, Fan Q, Kim DW, Kaufman HL. Local IL-21 promotes the therapeutic activity of effector T cells by decreasing regulatory T cells within the tumor microenvironment. Molecular Therapy. 2009;17(2):380–8.

21. Chesney J, Lewis KD, Kluger H, Hamid O, Whitman E, Thomas S, et al. Efficacy and safety of lifileucel, a one-time autologous tumor-infiltrating lymphocyte (TIL) cell therapy, in patients with advanced melanoma after progression on immune checkpoint inhibitors and targeted therapies: Pooled analysis of consecutive cohorts . J Immunother Cancer. 2022;10(12):1–14.

22. Wiltbank TB, Giordano GF, Kamel H, Tomasulo P, Custer B. Faint and prefaint reactions in whole-blood donors: An analysis of predonation measurements and their predictive value. Transfusion (Paris). 2008;48(9):1799–808.

23. Gattinoni L, Finkelstein SE, Klebanoff CA, Antony PA, Palmer DC, Spiess PJ, et al. Removal of homeostatic cytokine sinks by lymphodepletion enhances the efficacy of adoptively transferred tumor-specific CD8+ T cells. Journal of Experimental Medicine. 2005;202(7):907–12.

24. Geukes Foppen MH, Donia M, Svane IM, Haanen JBAG. Tumor-infiltrating lymphocytes for the treatment of metastatic cancer. Mol Oncol. 2015;9(10):1918–35.

25. Rohaan MW, Van Den Berg JH, Kvistborg P, Haanen JBAG. Adoptive transfer of tumor-infiltrating lymphocytes in melanoma: A viable treatment option. J Immunother Cancer. 2018;6(1):1–16.

26. Fernández VA, Martínez PB, Granhøj JS, Borch TH, Donia M, Svane IM. Biomarkers for response to TIL therapy : a comprehensive review. J Immunother Cancer. 2024;

27. Chesney J, Lewis KD, Kluger H, Hamid O, Whitman E, Thomas S, et al. Efficacy and safety of lifileucel, a one-time autologous tumor-infiltrating lymphocyte (TIL) cell therapy, in patients with advanced melanoma after progression on immune checkpoint inhibitors and targeted therapies: Pooled analysis of consecutive cohorts . J Immunother Cancer. 2022;10(12):1–14.

28. Medina T, Chesney JA, Whitman E, Kluger H, Thomas S, Sarnaik A, et al. Long-term efficacy and safety of lifileucel tumor-infiltrating lymphocyte (TIL) cell therapy in patients with advanced melanoma : A 4-year analysis of the C-144-01 study. 2023.

29. Matthaios D, Balgkouranidou I, Neanidis K, Sofis A, Romanidis K, Pappa A, et al. Revisiting Temozolomide ’ s role in solid tumors : Old is gold ? 2024;15.

30. Andersen R, Donia M, Ellebaek E, Borch TH, Kongsted P, Iversen TZ, et al. Long-Lasting complete responses in patients with metastatic melanoma after adoptive cell therapy with tumor-infiltrating lymphocytes and an attenuated il2 regimen. Clinical Cancer Research. 2016;22(15):3734–45.

31. Kristensen NP, Heeke C, Tvingsholm SA, Borch A, Draghi A, Crowther MD, et al. Neoantigen-reactive CD8+ T cells affect clinical outcome of adoptive cell therapy with tumor-infiltrating lymphocytes in melanoma. Journal of Clinical Investigation. 2022;132(2):1–16.

32. Rotte A, Frigault MJ, Ansari A, Gliner B, Heery C, Shah B. Dose-response correlation for CAR-T cells: A systematic review of clinical studies. J Immunother Cancer. 2022;10(12):1–11.

33. Stoltenborg Granhøj J, Rohaan M, Holz Borch T, Presti M, Nijenhuis C, van Zon M, et al. 8P Phenotypic characterization of infused tumor-infiltrating lymphocytes (TIL) correlates with response to adoptive cellular therapy (ACT) in patients with metastatic melanoma (MM). Immuno-Oncology and Technology. 2022;16:100113.

34. Rosenberg SA, Yang JC, Sherry RM, Kammula US, Hughes MS, Phan GQ, et al. Durable complete responses in heavily pretreated patients with metastatic melanoma using T-cell transfer immunotherapy. Clinical cancer research. 2011 Jul;17(13):4550–7.

35. Borch A, Carri I, Reynisson B, Alvarez HMG, Munk KK, Montemurro A, et al. IMPROVE: a feature model to predict neoepitope immunogenicity through broad-scale validation of T-cell recognition. Front Immunol. 2024;15(April):1–17.

36. Leko V, Rosenberg SA. Identifying and Targeting Human Tumor Antigens for T Cell-Based Immunotherapy of Solid Tumors. Cancer Cell. 2020;38(4):454–72.

37. Rohaan MW, Gomez-Eerland R, van den Berg JH, Geukes Foppen MH, van Zon M, Raud B, et al. MART-1 TCR gene-modified peripheral blood T cells for the treatment of metastatic melanoma: a phase I/IIa clinical trial. Immuno-Oncology and Technology. 2022;15(C):100089.

38. Johnson LA, Morgan RA, Dudley ME, Cassard L, Yang JC, Hughes MS, et al. Gene therapy with human and mouse T-cell receptors mediates cancer regression and targets normal tissues expressing cognate antigen. Blood. 2009;114(3):535–46.

39. Linette GP, Stadtmauer EA, Maus M V., Rapoport AP, Levine BL, Emery L, et al. Cardiovascular toxicity and titin cross-reactivity of affinity-enhanced T cells in myeloma and melanoma. Blood. 2013;122(6):863–71.

40. Morgan RA, Chinnasamy N, Abate-daga DD, Gros A, Robbins F, Zheng Z, et al. Cancer regression and neurologic toxicity following anti-MAGE-A3 TCR gene therapy Richard. J Immunother. 2014;36(2):133–51.

41. Xie N, Shen G, Gao W, Huang Z, Huang C, Fu L. Neoantigens: promising targets for cancer therapy. Signal Transduct Target Ther. 2023;8(1).

42. Dolton G, Rius C, Wall A, Szomolay B, Bianchi V, Galloway SAE, et al. Targeting of multiple tumor-associated antigens by individual T cell receptors during successful cancer immunotherapy. Cell. 2023;186(16):3333–3349.e27.

43. Andersen RS, Thrue CA, Junker N, Lyngaa R, Donia M, Ellebæk E, et al. Dissection of T-cell antigen specificity in human melanoma. Cancer Res. 2012 Apr;72(7):1642–50.

44. Kvistborg P, Shu CJ, Heemskerk B, Fankhauser M, Thrue CA, Toebes M, et al. TIL therapy broadens the tumor-reactive CD8+ T cell compartment in melanoma patients. Oncoimmunology. 2012;(July):409–18.

45. Borgers JSW, Lenkala D, Kohler V, Jackson EK, Linssen MD, Hymson S, et al. Personalized, autologous neoantigen-specific T cell therapy in metastatic melanoma: a phase 1 trial. Nat Med. 2025 Mar 1;

46. Hansen UK, Church CD, Simões AMC, Frej MS, Bentzen AK, Tvingsholm SA, et al. T antigen–specific CD8+ T cells associate with PD-1 blockade response in virus-positive Merkel cell carcinoma. Journal of Clinical Investigation. 2024;134(8).

