## Supplemental material for "Safety and feasibility of blood-derived Multiple Antigen-Specific Endogenous T cells (MASE-T) for metastatic melanoma"

### Supplementary Paragraph S1

**Inclusion criteria**

1. Age ≥ 18 ≤ 75
2. Progressive disease on or after anti-PD-1/anti-PD-L1 monotherapy or progressive disease on or after anti PD-1 plus anti-CTLA-4 therapy
3. The patient has histologically confirmed metastatic cutaneous melanoma. Patients with metastatic ocular/mucosal or other non-cutaneous melanoma cannot be included
4. The patient is HLA-A2 positive
5. ≥ 10 % of lymphocytes in peripheral blood are CD8+
6. At least one measurable parameter according to RECIST version 1.1 guidelines
7. ECOG performance status of 0 or 1
8. No significant toxicity from previous cancer treatments (CTC ≤ 1)
9. Women of childbearing potential: Negative serum pregnancy test and must use effective contraception. This applies from screening and until 6 months after treatment. Birth control pills, spiral, depot injection with gestagen, subdermal implantation, hormonal vaginal ring and transdermal depot patch are all considered effective contraceptives
10. Men with female partner of childbearing potential must use effective contraception from screening and until 6 months after treatment. Effective contraceptives are as described above for the female partner. In addition, documented vasectomy and sterility or double barrier contraception are considered effective contraceptives
11. Signed statement of consent after receiving oral and written study information
12. Willingness to participate in the planned treatment and follow-up and capable of handling
13. The patient has met the following haematological and biochemical criteria:

a) AST and ALT ≤2.5 X ULN or ≤5 X ULN with liver metastases
b) Serum total bilirubin ≤1.5 X ULN or direct bilirubin ≤ ULN for patient with total bilirubin level > 1.5 ULN
c) Serum creatinine ≤1.5 X ULN
d) ANC (Absolute Neutrophil Count) ≥1,000/mcL
e) Platelets ≥ 75,000 /mcL
f) Hemoglobin ≥ 9 g/dL or ≥ 5.6 mmol/L

**Exclusion criteria**

1. Another malignancy or concurrent malignancy unless disease-free for 3 years
2. Requirement for immunosuppressive doses of systemic corticosteroids (>10 mg/day prednisone or equivalent) or other immunosuppressive drugs within the last 3 weeks prior to screening
3. Prior treatment with adoptive transfer of Tumor Infiltrating T cells (TIL)
4. Grade 3-4 adverse events upon treatment with PD-1 checkpoint inhibitors (only phase B)
5. Patients who have any CNS lesion that is symptomatic, greater than 1 cm in diameter or show significant surrounding edema on MRI scan will not be eligible until they have been treated and demonstrated no clinical or radiologic CNS progression for at least 2 months. However, patients with subclinical brain metastasis with a maximum of 4 metastasis < 1 cm can be included.
6. The patient has any condition that will interfere with patient compliance or safety (including but not limited to psychiatric or substance abuse disorders)
7. The patient is pregnant or breastfeeding
8. The patient has an active infection requiring systemic therapy
9. The patient has received a live virus vaccine within 30 days of planned start of therapy
10. Significant medical disorder according to investigator; e.g severe asthma or chronic obstructive lung disease, dysregulated heart disease or dysregulated diabetes mellitus.
11. Concurrent treatment with other experimental drugs
12. Any significant active autoimmune disease
13. Severe allergy or anaphylactic reactions earlier in life
14. Known hypersensitivity to one of the active drugs or one or more of the excipients.
15. Unrelieved lower urinary tract obstruction

### Supplementary Table S1

| Protein | Sequence | Number of patients with reactivity (of n=87) | % of patients |
|---|---|---|---|
| MAGE-A2 | LVHFLLLKY | 27 | 31,03 |
| gp100 / Pmel17 | IMDQVPFSV | 14 | 16,09 |
| CDKN1A | GLGLPKLYL | 14 | 16,09 |
| gp100 / Pmel17 | YLEPGPVTA | 13 | 14,94 |
| Melan-A / MART-1 (WT) | EAAGIGILTV | 13 | 14,94 |
| MAGE-C2 | KVLEFLAKL | 12 | 13,79 |
| MAGE-A10 | SLLKFLAKV | 12 | 13,79 |
| gp100 / Pmel17 | KTWGQYWQV | 10 | 11,49 |
| STEAP1 | MIAVFLPIV | 10 | 11,49 |
| Telomerase | RLFFYRKSV | 10 | 11,49 |
| LAGE-1 | MLMAQEALAFL | 9 | 10,34 |
| MAGE-A2 | YLQLVFGIEV | 9 | 10,34 |
| MAGE-C2 | LLFGLALIEV | 9 | 10,34 |
| STAT1-alpha/ß | KLQELNYNL | 9 | 10,34 |
| TAG-1 | SLGWLFLLL | 9 | 10,34 |
| MC1R | TILLGIFFL | 8 | 9,20 |
| Melan-A / MART-1 | ILTVILGVL | 8 | 9,20 |
| TRP-2 | FVWLHYYSV | 8 | 9,20 |
| KIF20A | AQPDTAPLPV | 8 | 9,20 |
| MAGE-A10 | GLYDGMEHL | 7 | 8,05 |
| p53 | RMPEAAPPV | 7 | 8,05 |
| SSX-2 | KASEKIFYV | 7 | 8,05 |
| STEAP1 | FLYTLLREV | 7 | 8,05 |
| TRP-2 | SVYDFFVWL | 7 | 8,05 |
| GnTV | VLPDVFIRCV | 6 | 6,90 |
| Livin (ML-IAP) | SLGSPVLGL | 6 | 6,90 |
| MAGE-A1 | YLEYRQVPV | 6 | 6,90 |
| Meloe-1 | TLNDECWPA | 6 | 6,90 |
| NY-ESO-1 / LAGE-2 | SLLMWITQC | 6 | 6,90 |
| TRAG-3 | ILLRDAGLV | 6 | 6,90 |

Top 30 most frequently detected melanoma tumor-associated antigens (TAA) in a cohort of 87 metastatic melanoma patients screened for reactivity against ~160 A0201-restricted TAAs. Ag-scaffolds targeting T cells specific towards these 30 TAAs were used to expand the MASE-T product

### Supplementary Paragraph S2

#### A) pMHC and tetramer generation

The 30 selected melanoma TAA peptides (listed in Supplementary Table 1) were purchased from Pepscan (Pepscan Presto BV) and dissolved to 10mM in DMSO. A functionally empty disulfide-stabilized variant of HLA-A*02:01 monomer was loaded with each peptide by incubation for 30min. at room temperature (Saini et al. Sci Immunol. 2019). Exchanged pMHC were centrifuged for 5min. at 3300g to avoid unwanted aggregates. Tetramers were generated by addition of 1,804ug streptavidin-fluorochrome-conjugate per 100uL pMHC (100ug/mL) and incubation for 30min. at 4°C. Following this, D-Biotin (Avidity, BIO-200) was added at a final concentration of 25uM (Supplementary Table 2).

#### B) Ag-scaffold assembly

The 30 individual Ag-scaffolds included in the multi-targeting melanoma Ag-scaffold were generated at the Technical University of Denmark (DTU), Department of Health Technology, as previously described, combined and stored at -80°C until use (Tvingsholm et al. J Immunother Cancer, 2023 ). In short, Ag-scaffold assembly involves co-incubation of avi-tagged and biotinylated pMHC and cytokines (IL-2 Avitag; Acro IL-2-H82F3, IL-21 Avitag, Acro IL-21-H82F7) with streptavidin-conjugated dextran backbone (500kDa, Fina Biosolutions) (Figure 1A).

#### C) Flow cytometry-based bead assay for Ag-scaffold identity testing

To confirm that Ag-scaffolds retained structural integrity and maintained their display of pMHC, IL-2, and IL-21 after storage, we performed a flow cytometry-based identity assay. Ag-scaffolds were captured on fluorescent beads conjugated with anti-dextran antibodies (StemCell Technologies). Next, the captured Ag-scaffold were incubated with fluorescence-conjugated antibodies to detect each of the individual components (pMHC (antibody directed against b2-microglobulin), IL-2 and IL-21), followed by flow cytometry analysis (Supplementary Table 2)

### Supplementary Table S2

| Marker | Conjugate | Clone | Vendor | Cat no | | Marker | Conjugate | Clone | Vendor | Cat no |
|---|---|---|---|---|---|---|---|---|---|---|
| CD3 | FITC | SK7 | BD | 345764 | | PD-1 | BV421 | EH12.1 | BD | 565024 |
| CD8a | BV480 | RPA-T8 | BD | 566121 | | GZMb | Alexa700 | QA16A02 | Biolegend | 304753 |
| CD8a | BV510 | RPA-T8 | BD | 563256 | | CD39 | PE-CF594 | TU66 | BD | 563678 |
| CD8a | QD605 | SK1 | BD | 563919 | | TCF | PE | S33.966 | BD | 564217 |
| HLA-A02 | FITC | BB7.2 | Abcam | ab27728 | | Ki67 | BUV395 | B56 | BD | 564071 |
| | | | | | | CD28 | BUV737 | CD28.2 | BD | 612815 |
| IL-2 | PE | MQ1-17H12 | BD | 554566 | | CD127 | PE-CF594 | HIL-7R-M21 | BD | 562397 |
| IL-21 | PE | 3A3-N2.1 | BD | 560463 | | CD45RA | BV711 | HI100 | BD | 563733 |
| MHC | FITC | TU99 | BD | 551338 | | CCR7 | FITC | G043H7 | Biolegend | 353216 |
| anti-SA | PE | 3A20.2 | Biolegend | 410503 | | fox p3 | PE-CF594 | 259D/C7 | BD | 562421 |
| | | | | | | CD4 | BV650 | SK3 | BD | 563875 |
| CD107a | BV421 | H4A3 | BD | 562623 | | Tox | APC | REA473 | Milenyi Biotec | 5201005445 |
| TNF | APC | MAb11 | Biolegend | 502912 | | CD3 | BV786 | SK7 | BD | 349301 |
| IFNg | PE-Cy7 | x | BD | 557643 | | CD69 | BUV395 | FN50 | BD | 564364 |
| | | | | | | TCF | BV421 | S33-966 | BD | 566692 |
| SA | PE | | Biolegend | 405204 | | CD27 | BV605 | O323 | Biolegend | 338033 |
| SA | APC | | Biolegend | 405243 | | CD57 | FITC | HNK-1 | Biolegend | 359604 |
| SA | BV421 | | BD | 563259 | | CD39 | BV786 | TU66 | BD | 742523 |
| SA | PE-CF594 | | BD | 562284 | | LAG3 | FITC | 17B4 | Adipogen | AG-20B-0012F-C100 |
| SA | PE-Cy7 | | Biolegend | 405206 | | Tim3 | PE-Cy7 | F38-2E2 | Biolegend | 345014 |
| SA | BV650 | | BD | 563855 | | CD4 | PE-Cy7 | RPA-T4 | BD | 560649 |
| SA | BUV737 | | BD | 564293 | | CD8a | BUV395 | RPA-T8 | BD | 563795 |
| SA | BV786 | | BD | 563858 | | Ki67 | BV711 | B56 | BD | 563755 |
| SA | BUV395 | | BD | 564176 | | CCR7 | APC | G043H7 | Biolegend | 353214 |

**Suppl. Table S2 |** Conjugates and antibodies.

### Supplementary Paragraph S3

#### A) Phenotyping of MASE-T products and PBMCs

Phenotypic characterization of the MASE-T infusion products (range from 0.6 to $5*10^6$ cells per sample) and PBMCs (range from 0.15 to $2.5*10^6$ cells per sample) collected at day -5 and 1, 3, 6, 12, and 18 weeks post-MASE-T infusion was performed on cryopreserved material using flow cytometry, based on the antibody panels outlined in Supplementary Table S2. Furthermore, monitoring of TAA-responsive cells using tumor antigen HLA-A2 MHC class I tetramers (Supplementary Figure S1A) was performed as during the screening procedure.

#### B) Intracellular cytokine staining

Autologous tumor cell lines (TCLs) were expanded from 2-3 mm tumor pieces in RPMI 1640 (Gibco) with 10% fetal bovine serum (FCS, Thermo Fisher) and 1% penicillin-Streptomycin (Gibco). Tumor digest was prepared by enzymatic digestion of tumor tissue (BD dissociation kit, BD 661563). For IFNγ-stimulation of tumor cell lines, TCLs were preincubated in 100 IU/ml IFNγ(BD) for 72h prior to co-culture with T cells. T cells were thawed and rested in X-VIVO 15 supplemented with 5% HS for 24h prior to co-culture. T cells ($3*10^5$ cells/well) and tumor cells ($1*10^5$cells/well) were combined in a 96-well plate. The negative control ('Alone'), had no tumor cells and the positive control (PMA/Ion), was supplemented with 25 ng/mL PMA and 0.5 µM Ionomycin (Sigma-Aldrich). All co-cultures were topped up with Golgi solution containing anti-CD107a antibody (Supplementary Table S2)), GolgiStop (BD Biosciences) and GolgiPlug (BD Biosciences), to a final co-culture volume of 200 µL. Co-cultures were incubated at 37°C, 5% $CO_2$ for 8h before harvest. Subsequently the cells were stained with live/dead (NiR) marker followed by staining for surface markers (CD3, CD8, CD4, CD137 and CD107a) and fixation and permeabilization for 1 hour. Finally, the cells were washed and stained for intracellular markers (TNF, INFG) followed by flow cytometry analysis.

### Supplementary Paragraph S4

**Validation and stability of the Ag-scaffold pool supplied to generate the multiple antigen-specific endogenously derived T cells (MASE-T) product**

Tumor-specific T cells can be selectively expanded from patient PBMCs using the Ag-scaffold technology, comprising a dextran backbone with co-attached pMHCs, directing the Ag-scaffold to antigen-specific T cells, and cytokines, providing a fine-tuned stimulatory signal that drives activation and proliferation (Figure 1A). By supplementing the Ag-scaffold every 3-4 days, tumor-specific T cells can be selectively expanded from patient PBMCs during a 14-day culture in G-Rex system (Figure 1B). To target relevant tumor-specific T cell populations in melanoma, we assembled and combined 30 Ag-scaffolds, each carrying an HLA-A*02:01-restricted peptide derived from common melanoma TAAs (listed in Supplementary Table S1). Using the multi-targeting Ag-scaffold mixture, we could efficiently increase the frequency and number of TAA-specific T cells from PBMCs from three melanoma patients (MM1-3) (Supplementary Figure S1A). Detection of 30 TAA-specific T cell populations before and after Ag-scaffold expansion using combinatorial encoded, fluorescently labeled peptide-MHC tetramer staining demonstrated that the total TAA-specific population in MM1 comprised more than nine different TAA-specific T cell populations that were expanded simultaneously (supplementary Figures S1B and S1C) (Andersen et al., Nat Protoc. 2012). To test the stability of the Ag-scaffolds during long-term storage at -80°C, we co-assembled an Ag-scaffold targeting EBV-specific T cells using the same components as the clinical trial batch. On the day of assembly (Day 0), this reference Ag-scaffold efficiently increased the frequency of EBV-specific T cells in four healthy donors (HD1-4, Supplementary Figure S2A) and maintained its expansion capacity for at least three years, covering the duration of our trial (Supplementary Figure S2B). Additionally, we analyzed the composition of the Ag-scaffolds after three years of storage at -80°C, demonstrating the continuous display of pMHCs, IL-2, and IL-21 on the dextran backbone (Supplementary Figure S2C). In the current trial, MASE-T expansion was performed on PBMCs isolated from a blood draw (300ml) on day -14. On expansion day 9 (Day -5), patients with a TAA-specific T cell fold expansion >5 were admitted for lymphodepleting chemotherapy (Day -4, -3 and-2) in order to pre-condition them for MASE-T infusion on Day 0. After hospital discharge, patients were evaluated for clinical response at week six  (Figure 1C).

**Supplementary Figure S1**

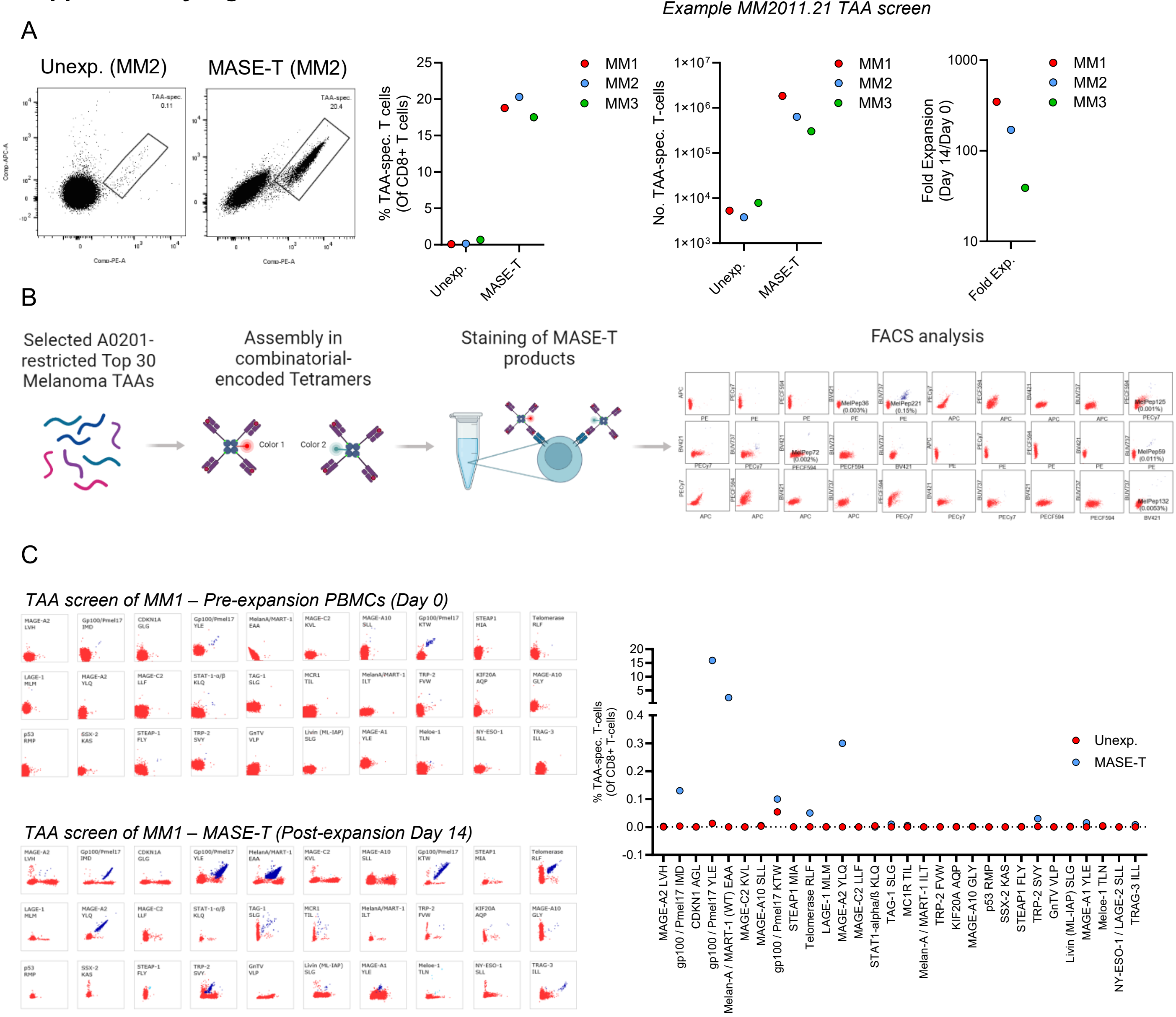


**Suppl. Figure S1 | Ag-scaffold-expansion of Top30 most frequently detected TAA responses in melanoma.**

**(A)** Example of Ag-scaffold mediated expansion of TAA-specific PBMCs from three metastatic melanoma-patients (MM1-3). The flow cytometry plots display TAA-specific cells before and after expansion, and the graphs show the frequencies, numbers, and fold expansion of TAA-specific cells. **(B)** Schematic drawing of parallel detection of TAA-specific T cell responses (n = 30) using combinatorial encoding with streptavidin-fluorochrome-based tetramers **(C)** Frequency of TAA-specific T cell populations in PBMCs from patient MM1 at baseline (Day 0) and after (Day 14) simultaneous expansion with scaffolds presenting the 30 TAAs. The graph on the right summarizes the identity and frequency of the responses detected in patient MM1 prior to and after expansion.

### Supplementary Figure S2

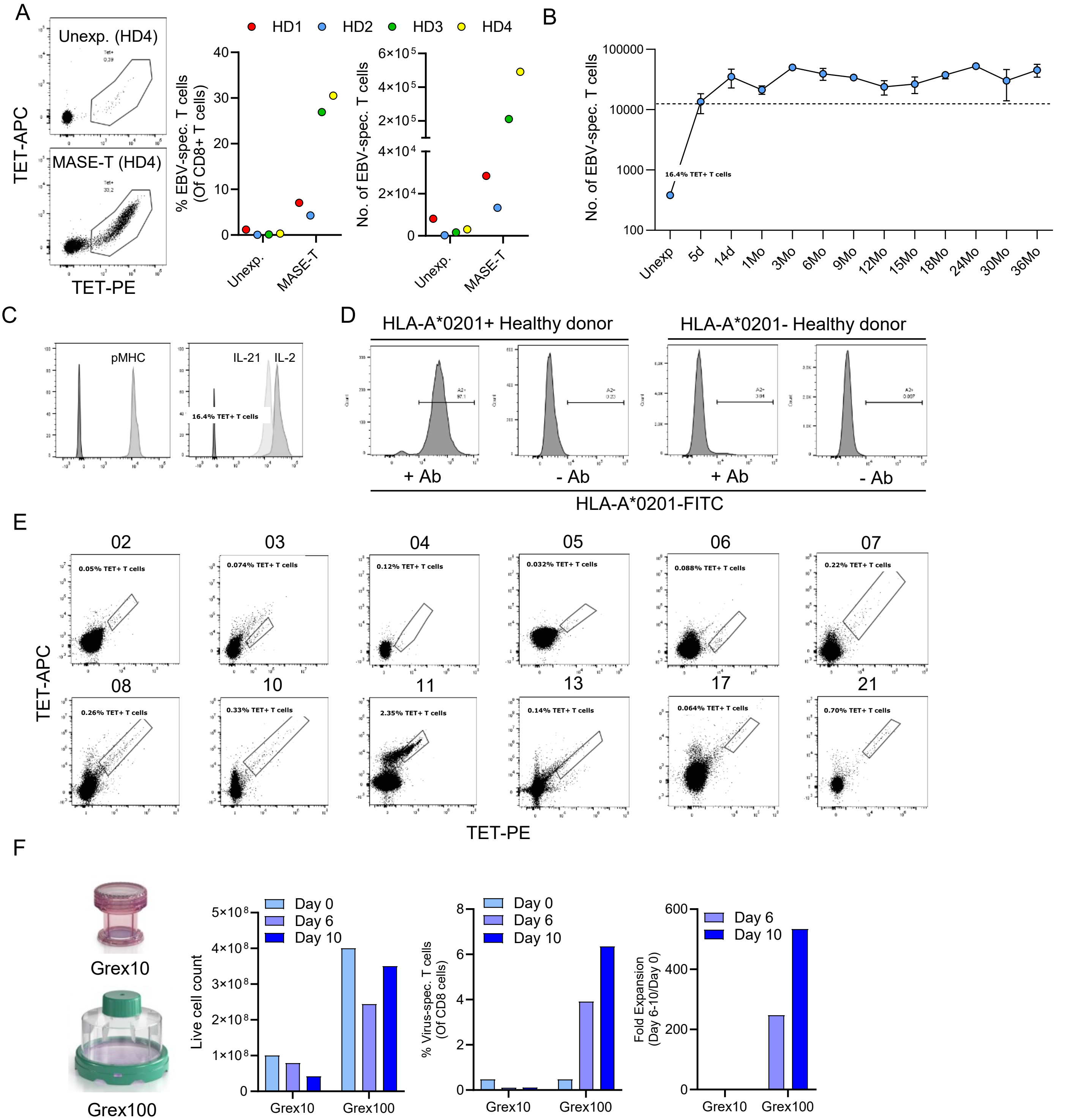


**Suppl. Figure S2 | (A)** Expansion of antigen-specific T cells from four healthy donors (HD1-4) by an EBV BMLF1-targeting Ag-scaffold, co-assembled in parallel with the multi-targeting Ag-scaffold used in the trial. The graphs display the frequencies and numbers of antigen-specific cells. **(B)** Evaluation of the capacity of the EBV BMLF1-targeting Ag-scaffold to expand EBV BMLF1-specific T cells from healthy donor PBMCs (HD2) following cryopreservation at -80°C for up to 3 years. **(C)** Anti-dextran bead-based flow cytometry assay to assess the presence of pMHC, IL-2 and IL-21 on the scaffold following three years of cryopreservation. **(D)** HLA-A*0201-typing of patient PBMCs by antibody-staining with a FITC-conjugated anti-HLA-A*0201 antibody. In HLA-A*0201-positive patients, the median FITC intensity is $>10^4$ upon antibody-staining (+ Ab) and remains $<10^4$ in an unstained control (- Ab). **(E)** PE-APC tetramer-staining for TAA-specific T cells (of CD8+ T cells) in PBMCs from the twelve patients that were included in the trial. The tetramer-positive gate is adjusted according to a positive control in each individual test. **(F)** Optimization of the Ag scaffold-expansion procedure after challenges experienced during MASE-T production in Grex10 for MM2011.02. The graphs compare total viable cell counts, and frequency and fold expansion of three combined virus-specific T cell populations (FLU M1 58-66, EBV BMLF1 and CMV pp65) from healthy donor PBMCs cells, on days 0, 6 and 10 of expansion in Grex10 and Grex100.

### Supplementary Figure S3

A

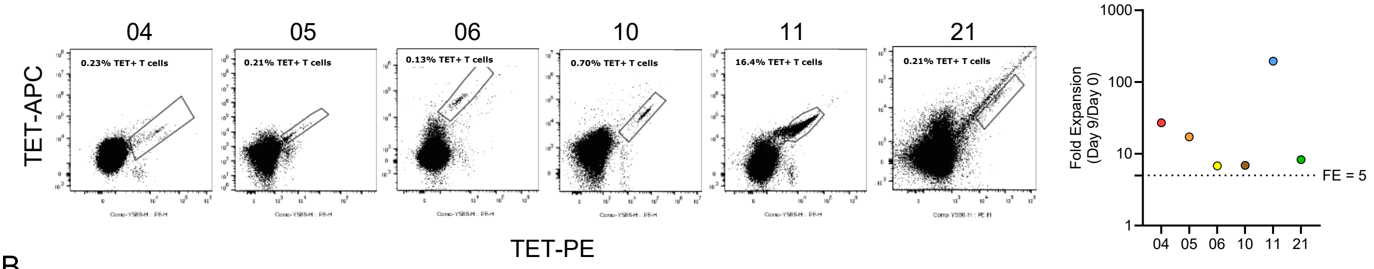


B

| Baseline characteristics | | | | | | | |
|---|---|---|---|---|---|---|---|
| **Patient** | **Sex** | **Histology** | **BRAF-mutated** | **Performance Status** | **Sites of Disease** | **Prior treatment lines** | **LDH baseline** |
| 04 | Male | Nodular melanoma | Yes | 0 | Liver, lungs, lymph nodes in the mediastinum, bones, CNS | Nivolumab, Ipilimumab, BRAF/MEK inhibitor, Temozolomide | 234 |
| 05 | Male | Nodular melanoma | No | 0 | Liver, lungs, lymph nodes in the mediastinum, Subcutis, CNS | Nivolumab, Ipilimumab/Nivolumab, PDL1-variant (E) | 189 |
| 06 | Female | Unclassified cutaneous melanoma | Yes | 0 | Small intestine, Intrapelvic and intraabdominal, axilla,subcutis | Pembrolizumab, BRAF/MEK inhibitor, Ipilimumab | 142 |
| 10 | Female | Unclassified cutaneous melanoma | No | 1 | Liver, multiple lymph nodes on the neck, subcutis | Pembrolizumab, Nivolumab+IDO vaccination (E), Ipilimumab, Temozolomide | 263 |
| 11 | Female | Unclassified cutaneous melanoma | Yes | 0 | Left adrenal gland, right mamma | Pembrolizumab, BRAF/MEK inhibitor, Ipilimumab, Radiotherapy, BRAF/MEK inhibitor | 312 |
| 21 | Female | Nodular melanoma | No | 0 | Lungs, retroperitoneum, axilla, subcutis | Pembrolizumab | 240 |

LDH = Lactate dehydrogenase

C D E

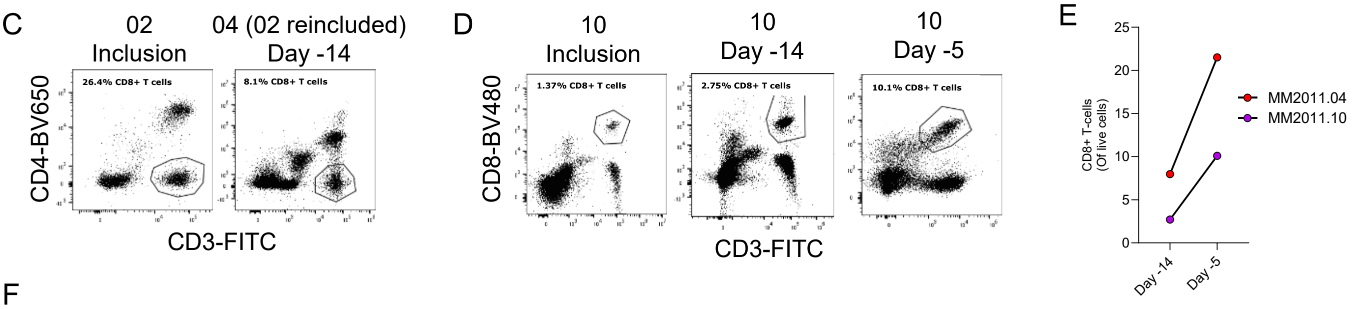


F

| Treatment after MASE-T | | |
|---|---|---|
| **Patient** | **Treatment post MASE-T** | **Best response** |
| 4 | BRAF/MEK inhibitor rechallenge | PD |
| 5 | Temozolomide | PD |
| 6 | BRAF/MEK inhibitor rechallenge, Tumor infiltrating lymphocytes | SD, PD |
| 10 | Radiotherapy | PD |
| 11 | Tumor infiltrating lymphocytes | PR |
| 21 | - | PD |

**Suppl. Figure S3 | (A)** PE-APC-tetramer-staining for TAA-specific T cells (of CD8+ T cells) from the six MASE-T treated patients on day 9 of expansion. The number of TAA-specific T cells on day 0 and 9 were used to calculate a fold expansion (FE), as displayed in the graph. **(B)** Extended baseline characteristics of the six MASE-T treated patients. **(C and D)** Frequency of **(C)** CD3+ CD4-, or **(D)** CD3+ CD8+ T cells of total lymphocytes at the inclusion, day -14 and day -5 PBMC samples from Patients **(C)** MM2011.02/04 and **(D)** MM2011.10. **(E)** Frequency of CD8+ T cells out of live cells in PBMC samples from patients MM2011.04 and 2011.10 on day -14 and day -5. **(F)** Treatments received after MASE-T therapy in the six treated patients and the best overall response obtained upon treatment.

**Supplementary Figure S4**

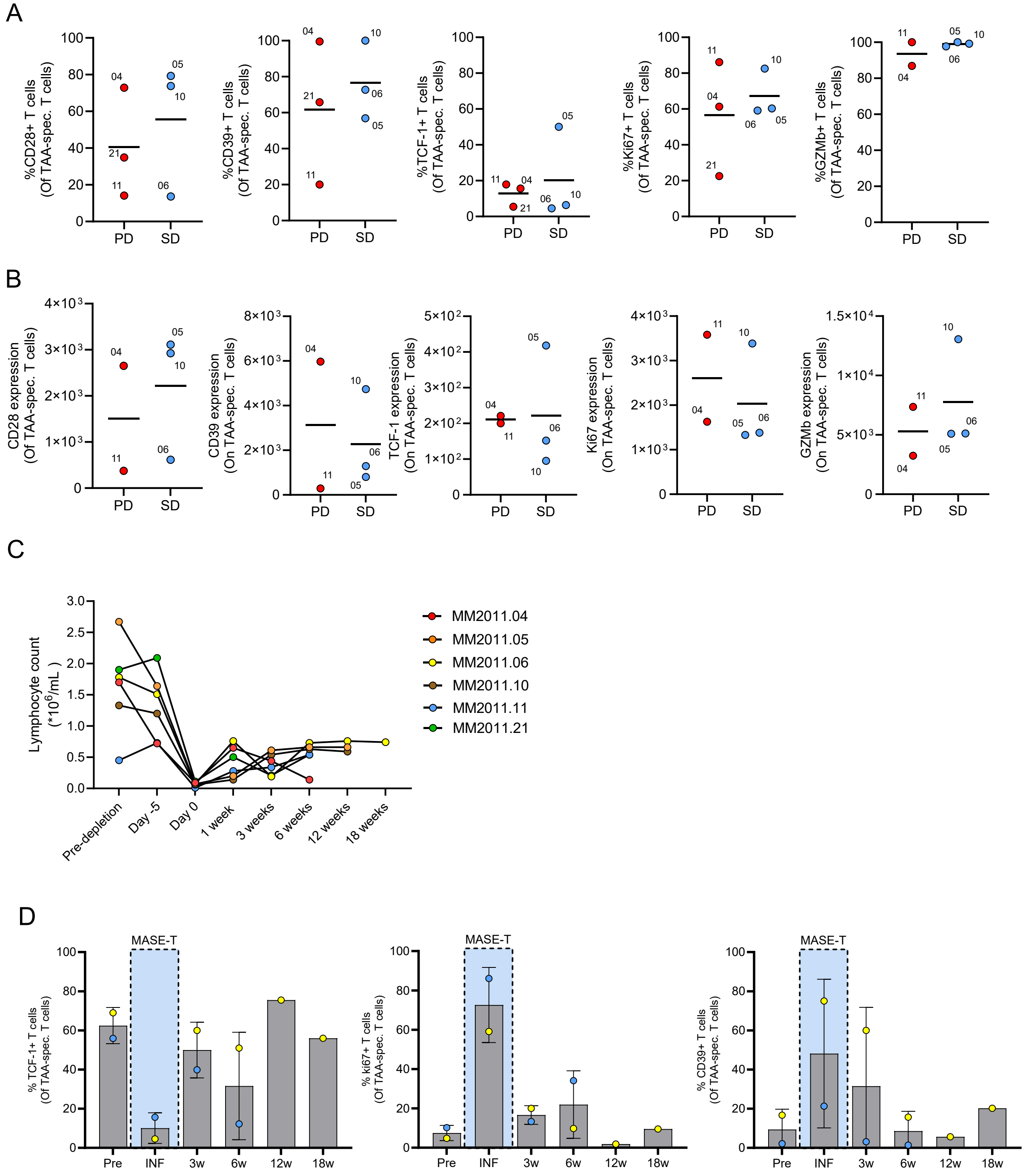


**Suppl. Figure S4 | (A)** Frequency and **(B)** MFI of TAA-specific cells expressing the indicated phenotypic markers in the cell products of MASE-T treated patients, following grouping according to the RECIST 1.1 classification of the patients. Horisontal lines represents the mean frequency/MFI. **(C)** Lymphocyte counts in PBMCs of MASE-T treated patients pre-lymphodepletion and at various time points after MASE-T treatment. **(D)** Frequency of expression of the phenotypic markers TCF-1 and CD39 and cell cycle-associated marked Ki67 on TAA-specific cells in the infused MASE-T products (INF), and in peripheral blood before (Pre) and at various time points after (3w, 6w, 12w and 18w) MASE-T treatment from patient MM2011.06 and MM2011.11 .

### Supplementary Figure S5

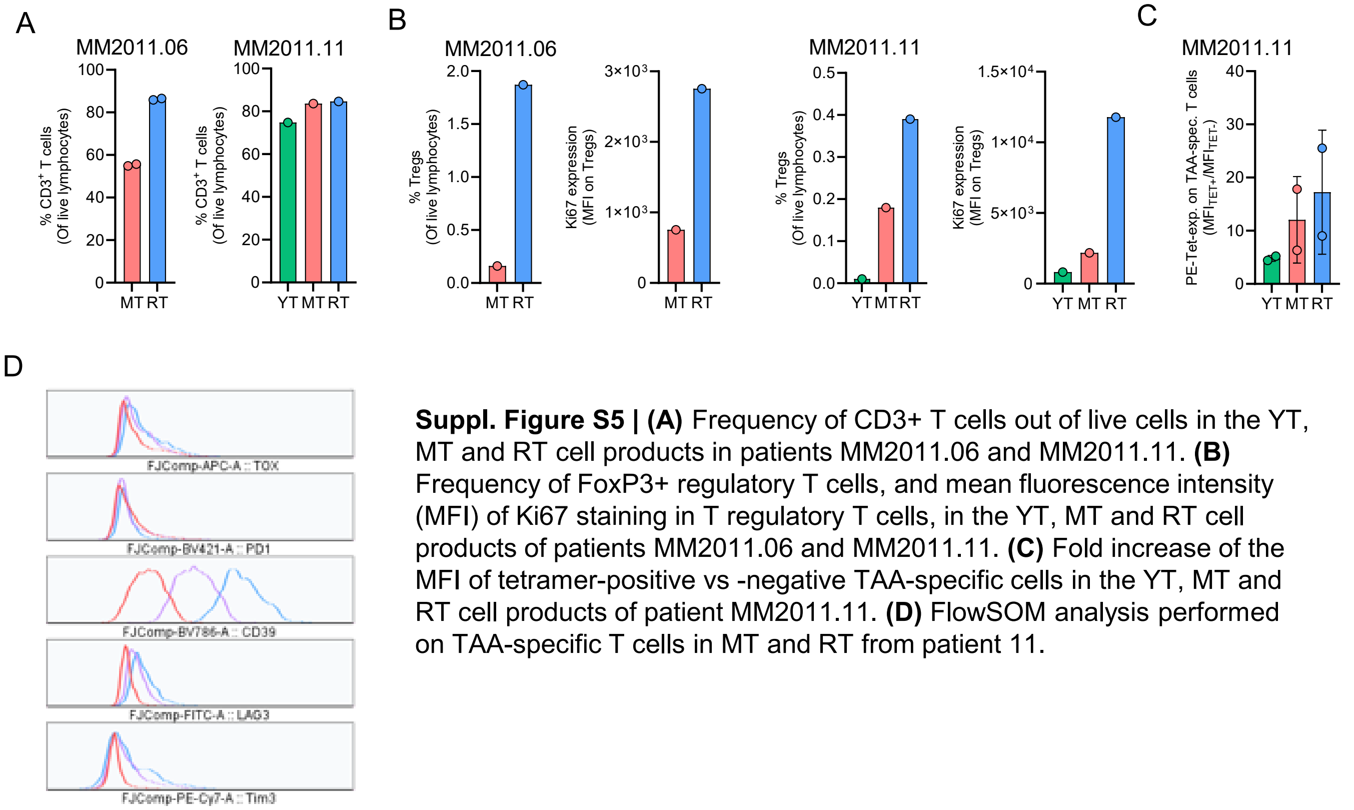


**Suppl. Figure S5 | (A)** Frequency of CD3+ T cells out of live cells in the YT, MT and RT cell products in patients MM2011.06 and MM2011.11. **(B)** Frequency of FoxP3+ regulatory T cells, and mean fluorescence intensity (MFI) of Ki67 staining in T regulatory T cells, in the YT, MT and RT cell products of patients MM2011.06 and MM2011.11. **(C)** Fold increase of the MFI of tetramer-positive vs -negative TAA-specific cells in the YT, MT and RT cell products of patient MM2011.11. **(D)** FlowSOM analysis performed on TAA-specific T cells in MT and RT from patient 11.
